# A Machine Learning Model for Post-Concussion Musculoskeletal Injury Risk in Collegiate Athletes

**DOI:** 10.1101/2025.01.29.25321362

**Authors:** Claudio C. Claros-Olivares, Melissa N. Anderson, Wei Qian, Austin J. Brockmeier, Thomas A. Buckley

## Abstract

**Background:** Emerging evidence indicates an elevated risk of post-concussion musculoskeletal (MSK) injuries in collegiate athletes; however, identifying athletes at highest risk remains to be elucidated.

**Objective:** The purpose of this study was to model post-concussion MSK injury risk in collegiate athletes by integrating a comprehensive set of variables by machine learning.

**Methods:** A risk model was developed and tested on a dataset of 194 athletes (155 in the training set and 39 in the test set) with 135 variables entered into the analysis, which included participant’s heath and athletic history, concussion injury and recovery specific criteria, and outcomes from a diverse array of concussions assessments. The machine learning approach involved transforming variables by the Weight of Evidence method, variable selection using L1-penalized logistic regression, model selection via the Akaike Information Criterion, and a final L2-regularized logistic regression fit.

**Results:** A model with 48 predictive variables yielded significant predictive performance of subsequent MSK injury with an area under the curve of 0.82. Top predictors included cognitive, balance, and reaction at Baseline and Acute timepoints. At a specified false positive rate of 6.67%, the model achieves a true positive rate (sensitivity) of 79% and a precision (positive predictive value) of 95% for identifying at-risk athletes via a well calibrated composite risk score.

**Conclusion:** These results support the development of a sensitive and specific injury risk model using standard data combined with a novel methodological approach that may allow clinicians to target high injury risk student-athletes. The development and refinement of predictive models, incorporating machine learning and utilizing comprehensive datasets, could lead to improved identification of high-risk athletes and allow for the implementation of targeted injury risk reduction strategies by identifying student-athletes most at risk for post-concussion MSK injury.

**Key Points:**

1. There is a well-established elevated risk of post-concussion subsequent musculoskeletal injury; however, prior efforts have failed to identify risk factors.
2. This study developed a composite risk score model with an AUC of 0.82 from common concussion clinical measures and participant demographics.
3. By identifying athletes at elevated risk, clinicians may be able to reduce injury risk through targeted injury risk reduction programs.

## 1 Introduction

The acute diagnosis of sports-related concussions has improved dramatically over the last two decades;[1–3] however, the determination of recovery remains an ongoing challenge.[4] Concussions affect cognitive function, motor control, vestibulocular and cardiovascular function and are associated with increased somatic and psychological symptoms.[1, 5–8] Current clinical assessments and neurological screening tools, while effective in identifying acute concussions, may not fully capture the persistent neurophysiological deficits that can linger beyond an athlete’s clinical recovery and return to participation (RTP).[4, 9, 10] The primary concern for post-concussion RTP decisions was the elevated risk of a subsequent concussion, but current concussion management protocols have significantly reduced this risk.[11] However, concerning evidence has emerged over the last decade of a two times elevated rate of musculoskeletal (MSK) injuries in the year following a concussion.[12–20] This relationship has been identified across diverse populations, including high school athletes,[16] collegiate athletes,[15, 17] military personnel,[18] and the general public.[19] These lower extremity MSK injuries pose substantial challenges to athletes acutely and may increase the rate of chronic conditions across the lifespan. Acutely, MSK injuries result in increased healthcare costs, lost school/work time, and elevated rates of mental health challenges.[21–24] Chronic conditions such as osteoarthritis, di”culty working, decreased quality of life, and elevated healthcare costs are more prevalent in former athletes with prior MSK injuries.[24–26] Thus, identifying athletes at risk for MSK injuries is a critical healthcare priority to improve athlete’s quality of life (QoL) and reduce healthcare costs.

Identifying athletes who are at an elevated risk for MSK injuries is possible but challenging due to the extensive set of potential modifiable and non-modifiable risk factors; indeed, a recent military MSK review identified 950 potential variables.[27–29] A systematic review of extensive previous efforts to develop MSK injury prediction models identifies some consistent risk factors, but results indicate generally limited success in overall prediction.[29, 30] The combination of relatively large samples and the number of potential variables has motivated machine learning/artificial intelligence models to improve outcome prediction.[30–33] A key limitation of these sports medicine injury prediction approaches is the lack of standard data sets.[34] Concussion studies and data sets offer an opportunity to overcome this limitation as assessment techniques generally follow international consensus guidelines while injury characteristics and athlete demographics are commonly recorded by healthcare providers.[4] As in the general MSK injury models, single or small groups of variables do not enable successful prediction models for post-concussion MSK.[17, 35] Thus, we propose a machine learning approach, consistent with military MSK injury risk approaches,[28] that incorporates an extensive set of risk factors, including 1) common clinical concussion assessments,[9] 2) demographic and anthropometric measures, 3) concussion injury characteristics, and 4) assessments during the post-concussion recovery process, as variables for a statistical model to predict subsequent MSK injury.

While using concussion assessments consistent with international guidelines will enable general adoption, challenges are still inherent with the use of a large set of heterogeneous variables measures collected at baseline and post-concussion. Three primary challenges are 1) missing data, 2) nonlinear relationships between the variables and the outcome, and 3) di”culties in encoding and combining categorical and continuous variables together without increases in dimensionality. To address these issues, we propose to use the weight of evidence (WoE) transformation to unify the set of variables before applying variable selection and logistic regression to model the risk of MSK.[36–40] WoE is a computationally e”cient variable transformation method that enhances the predictive power of variables, simplifies comparison across diverse data types, and curtails dimensionality growth. WoE has seen applications in credit scoring and risk assessment domains and may be well suited to address current limitations and challenges in identifying post-concussion MSK risk.[41, 42]

The elevated risk of MSK injury following a concussion is now well established; however, identifying accurate predictors of elevated risk has been unsuccessful. Therefore, the purpose of this study was to develop a machine learning-based post-concussion MSK risk model in collegiate athletes, with the composite risk score computed from commonly used clinical assessments as well as participant and injury characteristics. The successful development of this model may allow clinicians to implement targeted injury risk reduction approaches and reduce the post-concussion MSK incidence.

## 2 Methods

### 2.1 Participants

We recruited 211 National Collegiate Athletic Association (NCAA) student-athletes from a single institution who were diagnosed with a sports-related concussion. The inclusion criteria were any student-athlete who experienced a sports-related concussion and had at least one year of sports participation both before and after the concussion. Additionally, all participants completed a pre-career baseline assessment and completed the institution specific RTP protocol which was consistent with the contemporary consensus recommendations.[43, 44] The exclusion criteria were an athlete who did not return to participation following their concussion due to their athletic eligibility being completed, a decision to no longer participate following recovery (i.e., “quit”), or medical disqualification by the team physician.[45] Additional exclusion criteria included a concurrent injury with the concussion which required additional time loss beyond the concussion recovery (e.g., fracture). Missing time point(s) was not an exclusion criterion, but a participant had to have at least one time-point post-concussion to be included in this study. All participants provided written and oral informed consent as approved by the institution’s IRB (IRB Approval Numbers: 740790 and 804454, initial approval 2015). Out of the 211 athletes, 194 athletes met the inclusion criteria. (Table 1)

**Table 1:** Participants injury counts (a), demographics and anthropometrics (b) by grouped by Sex and Sport. No participant was engaged in more than one sport.

| Sport | Sex | Non-MSK<br>injury | MSK<br>injury | Event<br>rate |
| --- | --- | --- | --- | --- |
| Baseball/Softball | female | 4 | 8 | 0.667 |
|  | male | 4 | 3 | 0.429 |
| Basketball | female | 3 | 7 | 0.700 |
|  | male | 1 | 6 | 0.857 |
| Cheer | female | 3 | 3 | 0.500 |
|  | male | 1 | 1 | 0.500 |
| Field Hockey | female | 1 | 8 | 0.889 |
| Football | male | 10 | 18 | 0.643 |
| Lacrosse | female | 7 | 12 | 0.632 |
|  | male | 7 | 8 | 0.533 |
| Rowing | female | 6 | 5 | 0.455 |
| Soccer | female | 6 | 10 | 0.625 |
|  | male | 3 | 6 | 0.667 |
| Swimming and Diving | female | 7 | 3 | 0.300 |
|  | male | 2 | 0 | 0.000 |
| Tennis | female | 1 | 0 | 0.000 |
|  | male | 1 | 2 | 0.667 |
| Track and Field | female | 2 | 9 | 0.818 |
| Volleyball | female | 5 | 11 | 0.688 |

(a)
| Sex | Age@Baseline<br>(years) | Age@Concussion<br>(years) | Height<br>(cm) | Weight<br>(kg) |
| --- | --- | --- | --- | --- |
| male | 18.8±0.8 | 19.6±1.1 | 168.6±10.9 | 62.8±8.5 |
| female | 19.0±1.2 | 20.0±1.4 | 185.6±8.1 | 97.1±21.7 |

### 2.2 Instrumentation

Variables for the model included participant demographics and anthropometrics, concussion injury specific information, and a comprehensive concussion assessment at four times points: 1) pre-career ‘Baseline’ timepoint, and then at three additional post-injury time points, 2) Acute (*<*48 hours of injury), 3) Asymptomatic, and 4) RTP.[9]

Participant demographics included sex, prior concussion history (yes/no), total number of prior concussions (continuous variable), sport type (collision, contact, non-contact), and sport.[9] Additional variables included self-reported prior history of anxiety, ADD/ADHD, depression, learning disability, and other psychiatric disorder were recorded as binary variables (yes/no). Both the self-reported history of MSK injury (yes/no) and number of injuries (continuous) were recorded from the participants baseline health history for injuries prior to college following an IRB approved review of the athletic training electronic health record for MSK injuries during their collegiate careers.

The participants’ concussion specific information included loss of consciousness (yes/no) and post traumatic amnesia (yes/no) as binary variables. Additionally, both the days until asymptomatic and the days until RTP were included as continuous variables.

The concussion assessments included commonly used measures from both the SCAT as well as the NCAA/DoD CARE study, and the assessments were completed according to standard and well-established protocols.[9, 43, 44] All measures were performed at baseline and each of the three subsequent time points (acute, asymptomatic, RTP) with the exception of Hospital Anxiety and Depression Scale (HADS) and Satisfaction with Life Scale (SWLS) which were only performed at baseline.

The participant’s self-reported symptoms were recorded for both the number of symptoms endorsed (0–22) and the graded symptom (0–6) checklist (0–132), with a lower score reflecting fewer symptoms and severity.[43, 44]

The participants completed the balance error scoring system (BESS) which consists of three stances (double limb, single limb, and tandem) on two surfaces (firm, foam).[46–48] Deviations from the test position are considered errors with a maximum of 10 per condition for a total score range of 0–60 with a higher score reflecting worse balance. Participants also completed both a single and dual task tandem gait task.[49] Briefly, the tandem gait task requires the participants to walk heel-to-toe down a 3-meter line, turn, and return to the starting point. During dual task trails, participants responded to working memory cognitive challenges (e.g., subtraction by seven, spelling a 5-letter word backwards).[50] The outcome measure was the total time to complete each task with a higher time reflecting worse performance.[51, 52]

Cognition was assessed using the standard assessment of concussion (SAC),[43, 44] the ImPACT computerized cognitive assessment,[53, 54] and the Trail Making Test (TMT).[55] The SCAT-3 version of the SAC was used (i.e., 5-word memory list) with a possible 30 points and a higher score reflected better cognitive performance.[43, 44] The ImPACT neurocognitive assessment has four composite score outcome measures: verbal memory, visual memory, visual motor speed, and reaction time. For reaction time, a lower number reflects better performance as opposed to verbal memory, visual memory, and visual motor speed wherein a higher score reflects better performance. The TMT outcomes are the time taken to complete the task with a lower time reflecting better performance.

To assess vestibular and visual function, the participants completed the Vestibular Ocular Motor Screening (VOMS) and the King-Devick test.[56–59] The VOMS was scored as pass/fail based on an increase of more than 2 total symptom scores during the examination (smooth pursuits, horizontal and vertical saccades, horizontal and vertical ocular reflex, and visual motor sensitivity). The near point of convergence (NPC) was recorded as the mean of three trials and a lower score reflected a better NPC.[56–59] The King-Devick, performed either with spiral bound cards or a tablet, requires the participant to read three cards of random letters and the outcome measure is the total time to complete the task with a lower score reflecting better performance.[57, 58]

The participants completed eight trials of the clinical reaction time (CRT) test and the mean time of the trials was the outcome measure. A lower reaction time reflected better performance.[60, 61]

Clinical mental health screenings included the Brief Symptom Inventory-18 (BSI- 18), the Hospital Anxiety and Depression Scale (HADS), and the Satisfaction with Life Scale (SWLS).[9, 62–64] The BSI-18 is an 18-item self-reported questionnaire with a score range of 0–24, with higher scores being worse. The BSI has subsections including depression, anxiety, and somatization with specific thresholds for each: depression and anxiety greater than seven and somatization greater than six. The HADS score range is 0–21 with subsections for both anxiety and depression. The section specific threshold for each section is greater than eight. The SWLS as a 0–35 range with higher scores reflecting better life satisfaction and a score lower than 20 indicating low satisfaction.

### 2.3 Procedures

A concussion was initially identified by an athletic trainer and the diagnosis was confirmed by a team physician consistent with the current concussion consensus in sport group diagnostic criteria.[43, 44] The student-athletes were assessed for symptoms by an athletic trainer daily basis asymptomatic. Many, but not all, participants were enrolled as part of the NCAA-DoD CARE Consortium which influenced the postinjury testing timeline.[9] The concussion protocol required the student-athlete to be asymptomatic, achieve baseline or better values on the clinical exams, and complete a 6-day progressive return to activity protocol. Final clearance for unrestricted RTP was based on successful completion of the progressive exercise protocol and a normal physical examination performed by the Team Physician.

All participants were tracked for one-year post-RTP (e.g., a concussion RTP on October 1 would be tracked until September 30 of the following year) for sports related MSK injuries through the athletic training electronic medical records. An injury was defined as requiring treatment from the athletic training staff or team physicians and resulting in at least one day of limited activity.[17, 35]

### 2.4 Statistical Analysis

Our approach to form a risk model for MSK injury uses the Weight of Evidence (WoE) transformation, which is related to the naive Bayes classifier, combined with logistic regression analysis.[65, 66] Specifically, continuous and discrete variables are transformed by a binning approach, where the value assigned to each bin is the log-ratio of the frequencies of the variable falling into that bin under the two outcomes (MSK injury or not) and the bin edges are optimized in terms of their Information Value (IV) to enhance their predictive power (see Supplementary Material A.2 for more technical details). This value assignment quantifies the evidence in favor of the outcome to evidence against the outcome. Hence, the ‘weight of evidence’ denomination. It is worth noting that missing values can also follow this logic since they are still associated with an outcome, i.e, data imputation is handled automatically. Together, WoE’s optimal binning and transformation of values can model nonlinear relationships between the predictive variables and MSK injury risk, even in the presence of missing data. It should be noted that as a supervised data-driven technique, the WoE transformation is based solely on the training set.

In this study, each participant is represented by *P* = 135 predictive variables, which consider the 1) variables described in the Instrumentation section and 2) the difference between measures at baseline and all the other time points. For example, the difference in CRT between baseline and acute time points is calculated as Baselines minus Acute, and in the Figures is labeled `CRT Difference Baseline Acute`. Participants are randomly divided, with random numbers generation, into training and test sets (via stratified sampling to ensure a matched injury rate in both sets): the training set contains *N* = 155 athletes and the test set contains *N*_test_ = 39 athletes. The training set is comprised of 96 athletes who experienced an MSK injury after a concussion and 59 who did not. The test set contains 24 athletes who suffered a post-concussion MSK injury and 15 who did not. The test set is a held-out set that is only used for evaluation purposes and has no influence on the model training process.

The model formulation begins with denoting the MSK injury outcome variable as *Y* → {0, 1}, with *Y* = 1 for an athlete that has a subsequent MSK injury and *Y* = 0 for one without an injury. Let **x** = [*x*_1_,. .., *x*_*P*_ ] denote a vector of *P* predictive variables. The log-odds of an athlete having an MSK injury is defined as 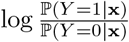, where ℙ(*Y* = 1|**x**) is the probability of an athlete having an MSK injury given the observed variables **x**.

Assuming the variables are independently contributing evidence, the log-odds can be expressed in terms of the WoE transformation applied to each variable (see Supplementary Material A). Combining all the variables, we fit a logistic regression model that weights each of the WoE-transformed variables as a refined estimate of the log-odds so that

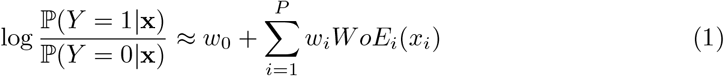

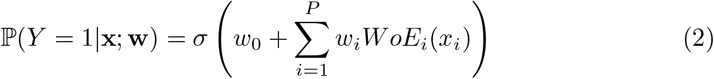

where ℙ(*Y* = 1|**x**; **w**) denotes the model’s probability estimate of MSK injury, 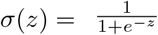 the logistic function, **w** = [*w*_0_,. .., *w*_*P*_ ] is the vector of parameters (bias and coe”cients), and *WoE*_*i*_(*x*_*i*_) denotes the transformation for the *i*th variable.

To remove any unnecessary variables from the model and enhance model performance by mitigating the issue of overfitting, we employ an L1-penalty when fitting the logistic regression model. The L1 penalty has the effect of forcing some coe”cient estimates to be exactly zero, effectively removing those variables from the model. The regularization parameter of the L1 penalty is systematically varied, creating a family of models with different numbers of selected variables and predictive performance. For model selection, we use the corrected Akaike Information Criterion (AICc), which balances model fit and complexity[67, 68] and provides asymptotically loss-e”cient variable selection when the true model is not contained within the candidate variable sets (a realistic assumption in many applications).[69] Finally, an L2-regularized logistic regression model is fit using the set of variables with non-zero coe”cients, with the regularization ameliorating any collinearity among the selected WoE-transformed variables. The performance of this model is evaluated on the test set by computing the area under the ROC curve (AUC) and area under the precision-recall curve, which is known as the average precision (AP) and shows the behavior of the accuracy of positive predictions made by a classifier (precision) against the classifier’s ability to identify all relevant instances within the positive class (recall).

As a benchmark, we also fit a logistic regression model which uses the original variables instead of the WoE-transformed variables. Mean imputation is applied when data is missing for this approach. Additionally, categorical variables are one-hot encoded and the remaining variables are standardized to have zero mean and unit variance so that their magnitudes can be comparable. (see Supplementary Material B)

After model fitting and selection, we apply model explanation techniques to identify the variables that contribute the most to the predictions. Specifically, we adopt Shapley Additive Explanations (SHAP)[62] which quantify the contribution of each variable to the prediction for each subject. In linear models, SHAP values are a product of the model’s coe”cients and the deviation of the variable from the mean. More specifically, for a given *x*, the contribution *ε*_*i*_ of the *i*th variable on the log-odds ratio estimate 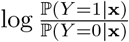 is

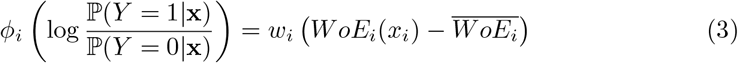

where 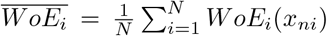 is the mean of the *i*th variable after the WoE transformation across the sample 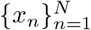 of size *N* . To summarize the impact of each variable, the absolute contributions are averaged by

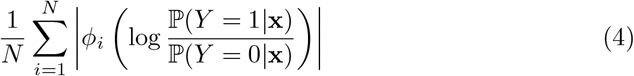

## 3 Results

Starting with *P* = 135variables that could be used to compose a risk, the AICc criterion determined that the simplest model that can explain the variance in the outcome variable only needs 48 variables (see Supplementary Appendix B for more details on the model selection and Supplementary Table B1 for a detailed list of the selected variables). The model trained with the selected variables was tested with data from athletes in the held-out test set. The distribution of log-odds ratios for the two groups of athletes (MSK injury and non-MSK injury groups) were significantly different (*p* = 0.003). (Figure 1)

**Fig. 1:**
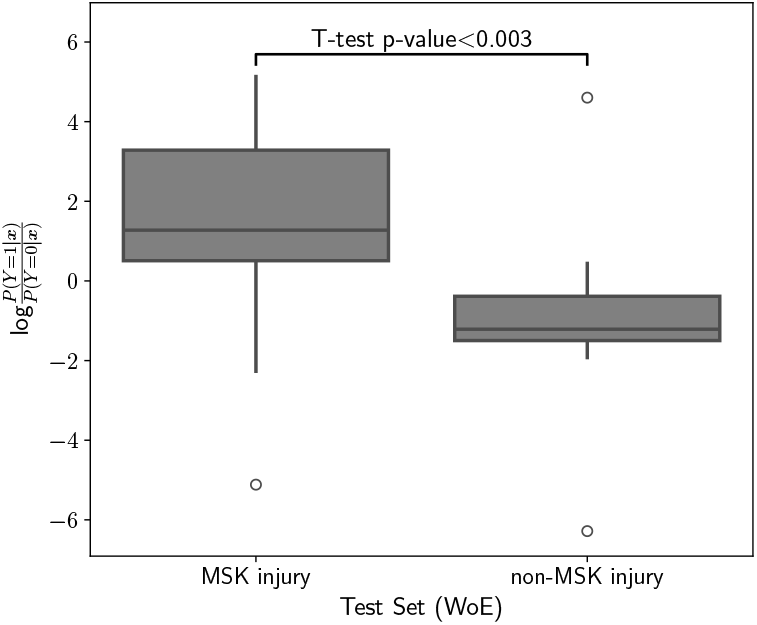
Distribution of Logit Scores for MSK injury and Non-MSK injury Groups in the Test Set. The box plot illustrate the spread and central tendency of the logit risk scores computed for our proposed approach. The model trained for WoE-transformed variables shows a statistical di!erence between the two groups. Q: Quartile

The ROC curve for the WoE-based model has an Area Under the Curve (AUC) of 0.82 which indicates a strong and clinically meaningful discriminatory ability of the model between the MSK and non-MSK groups. (Figure 2a). At a false positive of 6.67%, a true positive rate (recall/sensitivity) of 79% is achieved; this corresponds to a precision (positive predictive value) of 95%.

**Fig. 2:**
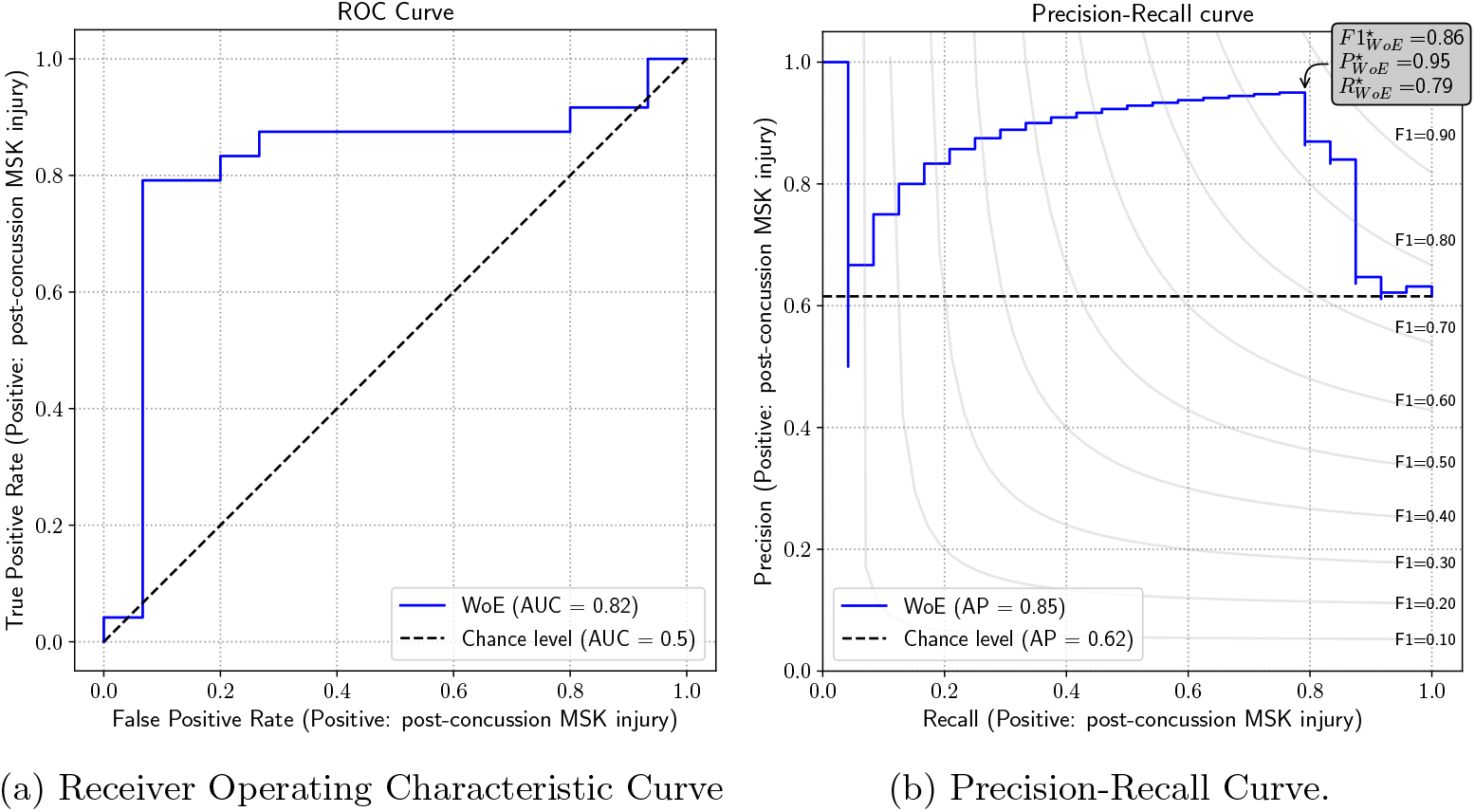
The ROC and Precision-Recall Curves for the Logistic Regression Models Post Variable Selection. The ROC curve (a) showcases the true positive rate (TPR) against the false positive rate (FPR) at various threshold levels with an overall AUC of 0.82. The precision-recall curve higlights (b) the precision (positive predictive value) at different levels of recall (sensitivity) with an average precision of 0.85. The best threshold would yield an F1 (harmonic mean of precision and recall) of 0.86.

The distribution of the probabilistic outputs for both the training and tests are provided in Figure 3. In the training set, (Figure 3a) the predictions indicate an overly optimistic performance, resulting from the model’s optimization to this specific dataset. The predictions for the test set exhibit a similar distribution to the training set, despite the model not having been exposed to this data previously. (Figure 3b) This similarity suggests that our model can generalize well to new data.

**Fig. 3:**
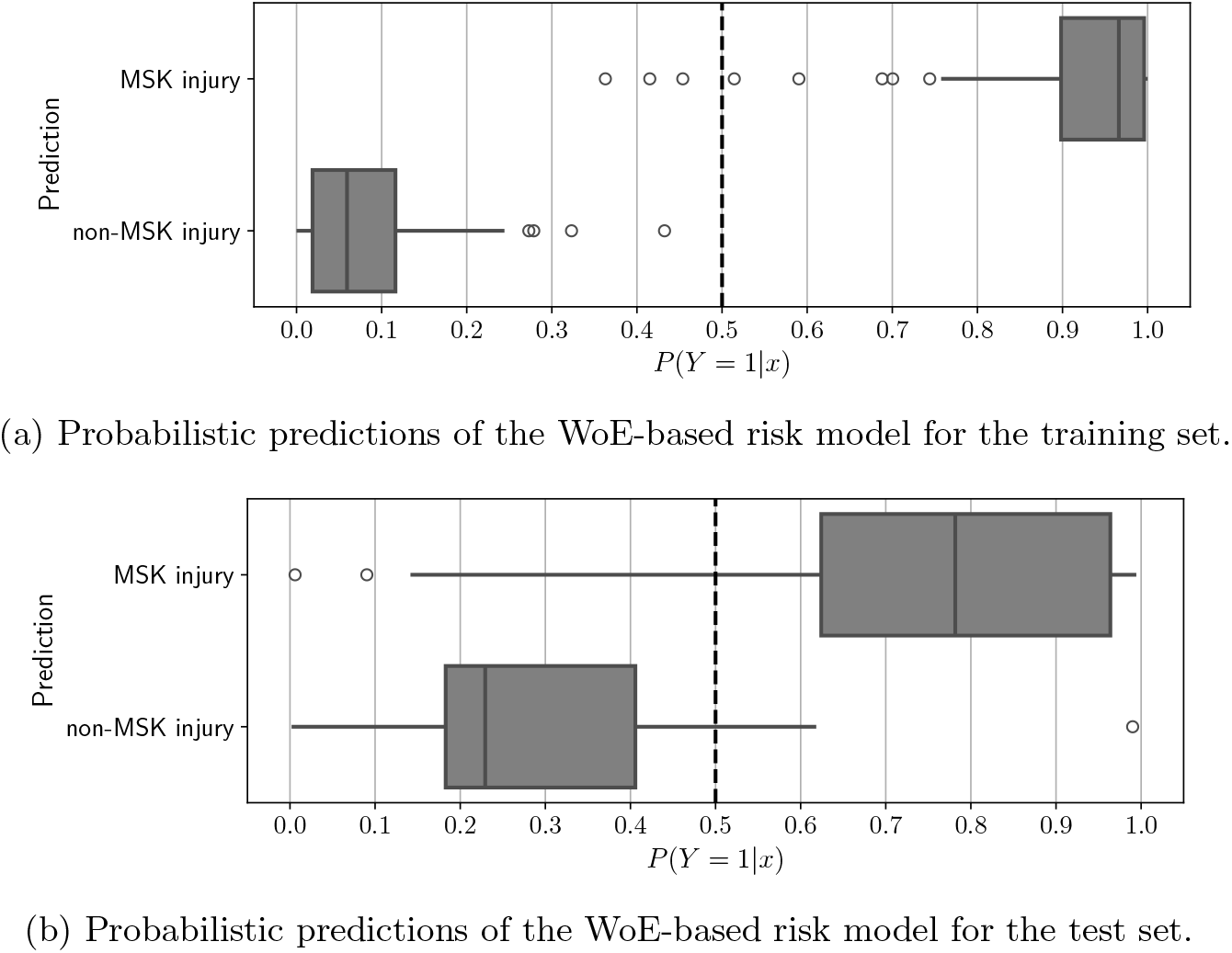
Probabilistic Predictions of a Logistic Regression Model for Injury Classification on (a) Training and (b) Test Sets. These plots illustrate the model’s predictions in probability space, differentiating post-concussion injury cases from non-injury ones. Notably, the behaviour of the predictions in the test set suggest that our trained model is able to make correct predictions for unseen data. Q: Quartile

Figure 4 presents the top 10 most influential variables in our trained logistic regression model. This ranking is based on the mean absolute SHAP values computed using the training set, which quantify each variable’s impact on the model’s predictions. No single variable or small group of variables dominates the predictions; rather, each variable contributes a small amount to the final outcome.

**Fig. 4:**
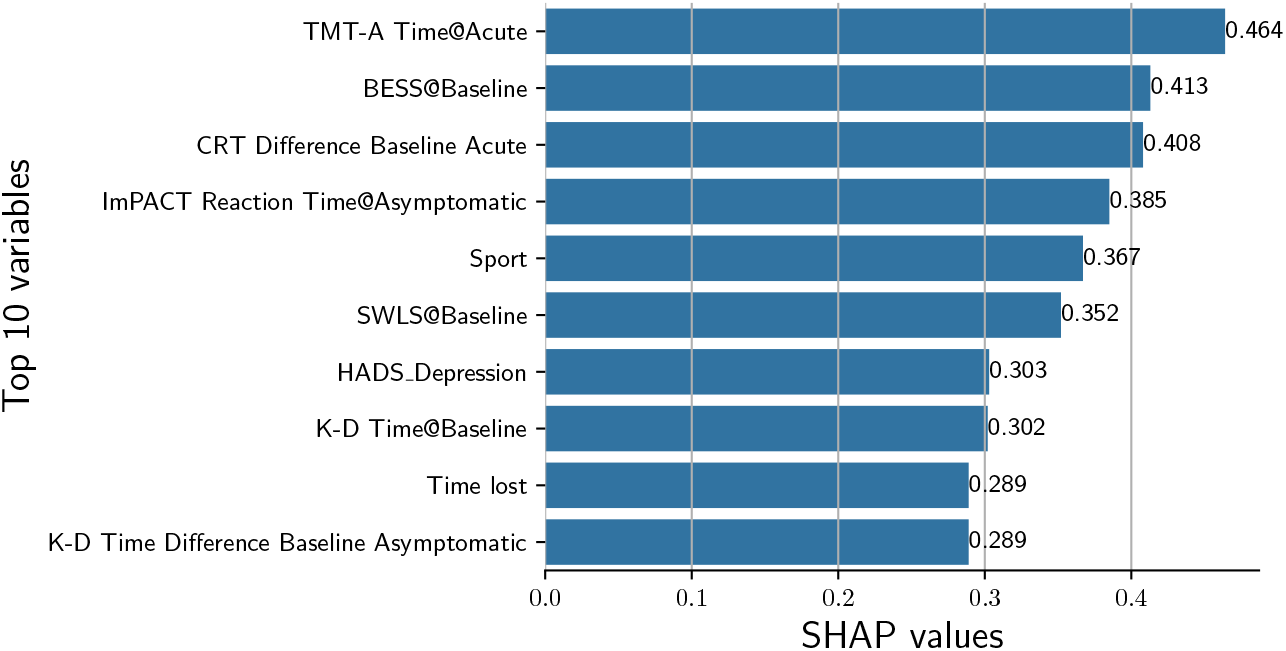
Top 10 Influential Variables in Logistic Regression Model Based on Mean Absolute SHAP Values. This bar chart shows the mean absolute SHAP values, computed from the training set, of the variables most influential to the model’s predictions. TMT-A: Trail Making Test-A; BESS: Balance Error Scoring System; CRT: Clinical Reaction Time test; ImPACT: Immediate Post-Concussion Assessment and Cognitive Test; SWLS: Satisfaction with Life Scale; K-D: King-Devick Test. Timepoints: Baseline – prior to athletic participation; Acute: within 48 hours of injury; Asymptomatic: when the patient reports no concussion related symptoms.

To assess the consistency of the proposed machine learning approach in producing a well-calibrated risk model with a subset of features, the methodology was applied to 20 random training-test splits of the dataset created using the stratified sampling scheme with the additional criterion that splits that resulted in a unique category for a categorical variable only appearing in the test set were rejected.

The results (Supplementary Material, Appendix C.2, Figures C5 and C6) from running the entire WoE transformation, feature selection, and model fitting selection showed a high degree of consistency with the model described above. Across the runs the mean and standard deviation of performance was AUROC: 0.79*±*0.05 (Supplementary Material, Table C6). The model selection criterion produced models with a similar number of features (average of 47) with a range of 41–52. Due to the correlation between many of the 135 variables (Supplementary Material, Figure C8), across the 20 runs a total of 112 distinct variables were selected and 31 of the 48 variables were in the top 50 variables most frequently selected. `Time Lost`, `Sport`, ImPACT Motor Speed@RTA, and TMT-A@Acute were included in all 20 runs (Supplementary Material, Figure C7). While the exact same variables were not chosen in each split, similar correlated variables were selected. The distribution of SHAP values shows that variables are consistent in their importance (Supplementary Material, Figure C9).

To assess whether the set of variables selected in the original split are as informative as other sets, we fit new models using these same variables across the 20 random train-test splits. The performance was nearly identical with AUROC: 0.80±0.03 (Supplementary Material, Table C6). This indicates the 48 variables are consistently good predictors of post-concussion MSK injury risk.

Additionally, we assessed the performance broken down by `Sport`. While predictive performance in terms of ROC curve is consistent for most sports (Supplementary Material, Figures C5 and C6), two sports: `Cheer` along with `Swimming and Diving` had notably lower performance, and two sports: `Tennis`, which had only 4 student-athletes in the study, and `Track and Field` had notably higher performance. Finally, we fit models without variables that have information on sport (Supplementary Material, Appendix C.5). Without these variables, performance is lower, but still fair, with AUC of 0.77± 0.05 (Supplementary Material, Table C7), which is not unexpected due to the varying prevalence of MSK injury across sports.

## 4 Discussion

In the year following a concussion, there is a well-established approximately two-fold elevated risk of a subsequent MSK injury.[12–19] In an ideal scenario, every athlete would receive personalized injury risk reduction treatments when returning to sports post-injury; however, this is neither logistically plausible nor cost-effective in amateur athletics. Due to the economic and personal costs associated with these injuries, a predictive risk algorithm that can identify athletes with higher risk can be used to identify those who may benefit from an injury risk reduction intervention. Previous attempts to develop predictive models using traditional analysis approaches considered individual predictors at single time points, but these attempts were largely unsuccessful.[17, 27, 35] Our study addresses this gap. We utilized a comprehensive set of variables based on commonly available injury information in a machine learning statistical modeling approach, which led to a well calibrated composite risk score for post-concussion MSK. The primary finding was the successful development of a clinically feasible model with high predictive accuracy (AUC: 0.82) to identify post-concussion MSK injury risk. This model may allow sports medicine clinicians to specifically target the highestrisk patients with established injury risk reduction programs.[70, 71]

An innovative component of our approach is the combination of WoE transformation and regularized logistic regression models with intrinsic feature selection to handle the extensive set of variables. Predictive models that use the WoE transformation are traditionally employed in the financial industry, particularly in credit scoring, risk assessment, and fraud detection. However, these methods have not been applied to model sports medicine injury risk challenges. Our approach combines WoE transformation with optimized binning of continuous and categorical variables as a data-driven approach to identify the best representation of the predictor variable.[36–40] By finding the binning scheme with the highest information value, the variable’s discriminatory power is maximized, leading to more accurate and robust predictive models. By integrating the WoE transformation with the feature selection capabilities of

L1-regularized logistic regression, our approach removes redundant predictors and mitigates model overfitting and ameliorates collinearity with subsequent L2-regularized logistic regression on the selected variables to achieve desirable risk segmentation even with a moderate sample size. Our approach also further addresses prior criticisms of MSK injury modeling by maintaining interpretable continuous variables, avoiding linearity assumptions, and using modeling techniques applicable to high dimensional data with various data types. Though the data set had 135 variables, the model developed herein uses only 48 variables (Supplemental Table B2), which can be reasonably included in concussion assessments and were all components of the NCAA-DoD CARE consortium assessment battery.[9] Furthermore, this is considerably less than the 950 potential MSK risk factors previously identified in military populations.[28, 29]

The variables contributing most to the model were a surprising mix of assessments and time points (Figure 4). The strongest predictors (in terms of the SHAP values) came from clinical assessments at both the baseline (pre-participation) and acute (¡48 hours) time points. A cornerstone of concussion management over the last decade is the use of a multifaceted assessment battery, and these results support this concept as the top predictor, which includes an array of cognitive, balance, and reaction time measures.[72–74] Each of these domains (i.e., cognition, oculomotor, and balance) has been associated with elevated injury risk, either conceptually or with limited empirical evidence, along with extrinsic factors such as sport and time loss. While each assessment technique has its limitations, especially concerning practice effects and testretest reliability,[75] taken together, they can create a composite score. Interestingly, SWLS was a highly influential variable in the predictive model. This was consistent with a prior report that found a 1-point increase in SWLS was associated with a 36% decrease in risk of MSK injury.[35] Lower satisfaction with life was associated with a higher risk of injury in Korean workers but has received limited application in sports medicine settings. Two of the top ten variables were not assessments but rather `Sport` and `Time lost`. Specific to sport, each sport had a different weight (Supplemental Table B2), which accounts for the inherent differences in the risk of MSK injury; future studies need to include a wider range of sports. Notably, athletes with nine or fewer days of time loss were at elevated risk of experiencing a post-concussion MSK, which is consistent with earlier studies showing elevated rates of subsequent concussion with short RTP durations.[11] A recent large CARE consortium finding (1,751 participants) reported the mean time to RTP was about 13 days, with only about 15% having full RTP within 7 days, suggesting that most athletes would not be within this risk category.[2] Taken together, this set of clinically feasible data points can be used to identify athletes at elevated risk of post-concussion MSK.

One inherent limitation of this model was the reliance on the influential effect of baseline data, which is a time and resource-intensive process.[76] Current consensus statements indicate that baseline data is not required to interpret assessments following a suspected concussion;[4] however, legal considerations (i.e., state laws) and compliance (e.g., Arrington settlement for NCAA member institutions) often dictate which baseline assessments will be performed.[77] As a large-scale project with numerous data collection time points and assessments, missing data was a clear limitation of the study (on average, roughly 35% was missing); nonetheless, a good predictive model was still developed. This model was developed from nearly 200 concussions at a single university and reproducibility of this model at other institutions, which may have varying distribution of athletes among sports and levels of competition, and other ages (e.g., high school, military) are critical steps forward. All data was collected by clinical athletic trainers or research staff well trained in the assessments, but inconsistency in data collection likely occurred. Another limitation of the model is its complexity. Future work could consider further simplifying the modeling by creating well-defined risk categories, similar to the military tra”c light system, to allow clinicians to apply risk reduction interventions to the highest-risk athletes.[28, 78] While the machine learning approach produced a particular model for the training set, similarly performing models with slightly different subsets of variables are produced on different splits (see Appendix C in the Supplemental Material). This is due to the relatively high correlation among some variables, as shown in a correlation heat map (Figure C8 in the Supplemental Material ). Relatedly, one drawback of the optimized binning of the WoE transformation is that bins created by the optimization algorithm on the training set may seem overly complicated. For example, the optimized bins for the variable `Time lost` (the number of days between concussion and RTP dates) generates a binning scheme with three segments: less than 9 days, between 9 and 12 days, and greater than 12 days. How best to simplify the model in a clinically meaningful manner, while retaining its predictive performance, requires further study. One possibility is to impose pre-determined bins when necessary for interpretation.

The results of this dataset analysis indicate that this approach holds the promise of providing robust and accurate risk categorization for post-concussion MSK injury. Incorporating more data from various institutions into the same methodology has the potential to produce an even stronger model. Moving forward, important next steps include a validation study to determine reproducibility and, if this is successful, developing a user-friendly and freely available online tool (e.g., website, app) to allow clinicians to input data to calculate the patient’s injury risk profile. By refining the model with additional data from multiple institutions and/or settings, we expect to enhance further its precision and applicability for clinicians to target high-risk post-concussion athletes with informed decisions, ultimately leading to more effective risk reduction strategies and programs to prevent post-concussion injury occurrences for high-risk athletes.

## 5 Conclusion

We proposed and developed an integrative analysis approach that combines numerous clinical measurement variables to assess the risk of subsequent MSK injury in a population of concussed athletes. The approach transforms variables by the WoE transformation before fitting a logistic regression model with variable selection to create a robust statistical model. The results from the risk score distribution and ROC curve analyses a”rm the resulting model is highly predictive of subsequent MSK injury, with sensitivity of 79% at a false positive rate of 6.67%, thus indicating the potential utility in practice.

## 6 Declarations

### Consent to Participate

Informed consent was obtained from all individual participants included in the study. There are no participants images or personally identifiable information in the manuscript.

### Authors Contributions

TB, AB, and WQ were involved in the study concept and design, interpretation of the data, statistical interpretation, manuscript drafting, and final approval. TB and MA were involved in the collection of the data. MA and CC were involved in the data analysis, interpretation of the data, statistical interpretation, manuscript drafting, and final approval. All authors read and approved the final version.

### Ethics Approval

The study was performed in accordance with the standards of ethics outlined in the Declaration of Helsinki. All study procedures were reviewed and approved by the University of Michigan IRB, the US Army Medical Research and Materiel Command Human Research Protection O”ce (HRPO) for all data related to the NCAA-DoD CARE Consortium. The University of Delaware Institutional Review Board approved all aspects of this study. IRB Approval Numbers: 740790 and 804454, initial approval 2015.

### Data Availability

The CARE Consortium datasets generated and analyzed during the current study are available in the FITBIR repository (https://fitbir.nih.gov/).

### Code Availability

Custom data analysis code written in Python with standard libraries is available on Github at https://github.com/cesar-claros/R21_risk_model/.

### Consent for Publication

Not Applicable.

## References

[1] Broglio, S.P., Harezlak, J., Katz, B., Zhao, S., McAllister, T., McCrea, M.: Acute sport concussion assessment optimization: a prospective assessment from the care consortium. Sports Medicine 49, 1977–1987 (2019)

[2] Broglio, S.P., McAllister, T., Katz, B.P., LaPradd, M., Zhou, W., McCrea, M.A.: The natural history of sport-related concussion in collegiate athletes: findings from the ncaa-dod care consortium. Sports Medicine 52(2), 403–415 (2022)

[3] Garcia, G.-G.P., Yang, J., Lavieri, M.S., McAllister, T.W., McCrea, M.A., Broglio, S.P., Investigators, C.C., et al.: Optimizing components of the sport concussion assessment tool for acute concussion assessment. Neurosurgery 87(5), 971–981 (2020)

[4] Patricios, J.S., Schneider, K.J., Dvorak, J., Ahmed, O.H., Blauwet, C., Cantu, R.C., Davis, G.A., Echemendia, R.J., Makdissi, M., McNamee, M., et al.: Consensus statement on concussion in sport: the 6th international conference on concussion in sport–amsterdam, october 2022. British Journal of Sports Medicine 57(11), 695–711 (2023)

[5] Dobson, J.L., Yarbrough, M.B., Perez, J., Evans, K., Buckley, T.: Sport-related concussion induces transient cardiovascular autonomic dysfunction. American Journal of Physiology-Regulatory, Integrative and Comparative Physiology 312(4), 575–584 (2017)

[6] Turner, S., Langdon, J., Shaver, G., Graham, V., Naugle, K., Buckley, T.: Comparison of psychological response between concussion and musculoskeletal injury in collegiate athletes. Sport, Exercise, and Performance Psychology 6(3), 277 (2017)

[7] Kontos, A.P., Eagle, S.R., Marchetti, G., Sinnott, A., Mucha, A., Port, N., Ferris, L.M., Elbin, R., Clugston, J.R., Ortega, J., et al.: Discriminative validity of vestibular ocular motor screening in identifying concussion among collegiate athletes: A national collegiate athletic association–department of defense concussion assessment, research, and education consortium study. The American Journal of Sports Medicine 49(8), 2211–2217 (2021)

[8] Buckley, T.A., Oldham, J.R., Caccese, J.B.: Postural control deficits identify lingering post-concussion neurological deficits. Journal of Sport and Health Science 5(1), 61–69 (2016)

[9] Broglio, S.P., McCrea, M., McAllister, T., Harezlak, J., Katz, B., Hack, D., Hainline, B.: A national study on the effects of concussion in collegiate athletes and us military service academy members: the ncaa–dod concussion assessment, research and education (care) consortium structure and methods. Sports medicine 47, 1437–1451 (2017)

[10] Kamins, J., Bigler, E., Covassin, T., Henry, L., Kemp, S., Leddy, J.J., Mayer, A., McCrea, M., Prins, M., Schneider, K.J., et al.: What is the physiological time to recovery after concussion? a systematic review. British journal of sports medicine 51(12), 935–940 (2017)

[11] McCrea, M., Broglio, S., McAllister, T., Zhou, W., Zhao, S., Katz, B., Kudela, M., Harezlak, J., Nelson, L., Meier, T., et al.: Return to play and risk of repeat concussion in collegiate football players: comparative analysis from the ncaa concussion study (1999–2001) and care consortium (2014–2017). British Journal of Sports Medicine (2019)

[12] McPherson, A.L., Nagai, T., Webster, K.E., Hewett, T.E.: Musculoskeletal injury risk after sport-related concussion: a systematic review and meta-analysis. The American journal of sports medicine 47(7), 1754–1762 (2019)

[13] Howell, D.R., Lynall, R.C., Buckley, T.A., Herman, D.C.: Neuromuscular control deficits and the risk of subsequent injury after a concussion: a scoping review. Sports Medicine 48(5), 1097–1115 (2018)

[14] Reneker, J.C., Babl, R., Flowers, M.M.: History of concussion and risk of subsequent injury in athletes and service members: a systematic review and meta-analysis. Musculoskeletal Science and Practice 42, 173–185 (2019)

[15] Brooks, M.A., Peterson, K., Biese, K., Sanfilippo, J., Heiderscheit, B.C., Bell, D.R.: Concussion increases odds of sustaining a lower extremity musculoskeletal injury after return to play among collegiate athletes. The American journal of sports medicine 44(3), 742–747 (2016)

[16] Lynall, R.C., Mauntel, T.C., Pohlig, R.T., Kerr, Z.Y., Dompier, T.P., Hall, E.E., Buckley, T.A.: Lower extremity musculoskeletal injury risk after concussion recovery in high school athletes. Journal of Athletic Training 52(11), 1028–1034 (2017)

[17] Buckley, T.A., Howard, C.M., Oldham, J.R., Lynall, R.C., Swanik, C.B., Getchell, N.: No clinical predictors of postconcussion musculoskeletal injury in college athletes. Medicine and Science in Sports and Exercise 52(6), 1256 (2020)

[18] Kardouni, J.R., Shing, T.L., McKinnon, C.J., Scofield, D.E., Proctor, S.P.: Risk for lower extremity injury after concussion: a matched cohort study in soldiers. Journal of Orthopaedic & Sports Physical Therapy 48(7), 533–540 (2018)

[19] McPherson, A.L., Shirley, M.B., Schilaty, N.D., Larson, D.R., Hewett, T.E.: Effect of a concussion on anterior cruciate ligament injury risk in a general population. Sports Medicine 50(6), 1203–1210 (2020)

[20] Hunzinger, K.J., Costantini, K.M., Swanik, C.B., Buckley, T.A.: Diagnosed concussion is associated with increased risk for upper extremity injury in community rugby players in males only. The Physician and Sportsmedicine (just-accepted) (2024)

[21] Abernethy, L., MacAuley, D.: Impact of school sports injury. British Journal of Sports Medicine 37(4), 354–355 (2003)

[22] Jayanthi, N.A., Post, E.G., Laury, T.C., Fabricant, P.D.: Health consequences of youth sport specialization. Journal of Athletic Training 54(10), 1040–1049 (2019)

[23] Knowles, S.B., Marshall, S.W., Miller, T., Spicer, R., Bowling, J.M., Loomis, D., Millikan, R., Yang, J., Mueller, F.: Cost of injuries from a prospective cohort study of north carolina high school athletes. Injury Prevention 13(6), 416–421 (2007)

[24] Larsen, E., Jensen, P., Jensen, P.: Long-term outcome of knee and ankle injuries in elite football. Scandinavian Journal of Medicine & Science in Sports 9(5), 285–289 (1999)

[25] Wikstrom, E.A., Hubbard-Turner, T., McKeon, P.O.: Understanding and treating lateral ankle sprains and their consequences: a constraints-based approach. Sports Medicine 43, 385–393 (2013)

[26] Wang, L.-J., Zeng, N., Yan, Z.-P., Li, J.-T., Ni, G.-X.: Post-traumatic osteoarthritis following acl injury. Arthritis Research & Therapy 22(1), 1–8 (2020)

[27] Oldham, J.R., Howell, D.R., Knight, C.A., Crenshaw, J.R., Buckley, T.A.: Gait performance is associated with subsequent lower extremity injury following concussion. Medicine and Science in Sports and Exercise 52(11), 2279–2285 (2020)

[28] Roach, M.H., Bird, M.B., Helton, M.S., Mauntel, T.C.: Musculoskeletal injury risk stratification: a tra”c light system for military service members. In: Healthcare, vol. 11, p. 1675 (2023). MDPI

[29] Rhon, D.I., Molloy, J.M., Monnier, A., Hando, B.R., Newman, P.M.: Much work remains to reach consensus on musculoskeletal injury risk in military service members: a systematic review with meta-analysis. European Journal of Sport Science 22(1), 16–34 (2022)

[30] Bahr, R.: Why screening tests to predict injury do not work—and probably never will…: a critical review. British journal of Sports Medicine (2016)

[31] Nassis, G., Verhagen, E., Brito, J., Figueiredo, P., Krustrup, P.: A review of machine learning applications in soccer with an emphasis on injury risk. Biology of sport 40(1), 233–239 (2023)

[32] Amendolara, A., Pfister, D., Settelmayer, M., Shah, M., Wu, V., Donnelly, S., Johnston, B., Peterson, R., Sant, D., Kriak, J., et al.: An overview of machine learning applications in sports injury prediction. Cureus 15(9) (2023)

[33] Bullock, G.S., Mylott, J., Hughes, T., Nicholson, K.F., Riley, R.D., Collins, G.S.: Just how confident can we be in predicting sports injuries? a systematic review of the methodological conduct and performance of existing musculoskeletal injury prediction models in sport. Sports medicine 52(10), 2469–2482 (2022)

[34] Kumar, G.S., Kumar, M.D., Reddy, S.V.R., Kumari, B.S., Reddy, C.R.: Injury prediction in sports using artificial intelligence applications: A brief review. Journal of Robotics and Control (JRC) 5(1), 16–26 (2024)

[35] Buckley, T.A., Bryk, K.N., Enrique, A.L., Kaminski, T.W., Hunzinger, K.J., Oldham, J.R.: Clinical mental health measures and prediction of postconcussion musculoskeletal injury. Journal of athletic training 58(5), 401–407 (2023)

[36] Siddiqi, N.: Credit Risk Scorecards: Developing and Implementing Intelligent Credit Scoring vol. 3. John Wiley & Sons, Hoboken, New Jersey (2012)

[37] Good, I.J.: Probability and the Weighing of Evidence. Charles Gri”n & Co. Ltd., London, UK (1950)

[38] Good, I.: Weight of evidence: a brief survey Bayesian Statistics 2 ed JM Bernardo, MH DeGroot, DV Lindley and AFM Smith. New York: Elsevier (1985)

[39] Singer, D.A., Kouda, R.: A comparison of the weights-of-evidence method and probabilistic neural networks. Natural Resources Research 8, 287–298 (1999)

[40] Hughes, G.: Youden’s index and the weight of evidence. Methods of information in medicine 54(02), 198–199 (2015)

[41] Anderson, R.: The Credit Scoring Toolkit: Theory and Practice for Retail Credit Risk Management and Decision Automation. Oxford University Press, Oxford, UK (2007)

[42] Thomas, L., Crook, J., Edelman, D.: Credit Scoring and Its Applications. SIAM, Philadelphia, Pennsylvania (2017)

[43] McCrory, P., Meeuwisse, W.H., Aubry, M., Cantu, R.C., Dvořák, J., Echemendia, R.J., Engebretsen, L., Johnston, K., Kutcher, J.S., Raftery, M., et al.: Consensus statement on concussion in sport: the 4th international conference on concussion in sport, zurich, november 2012. Journal of athletic training 48(4), 554–575 (2013)

[44] McCrory, P., Meeuwisse, W., Dvorak, J., Aubry, M., Bailes, J., Broglio, S., Cantu, R.C., Cassidy, D., Echemendia, R.J., Castellani, R.J., et al.: Consensus statement on concussion in sport—the 5th international conference on concussion in sport held in berlin, october 2016. British journal of sports medicine 51(11), 838–847 (2017)

[45] Schmidt, J.D., Rawlins, M.L.W., Lynall, R.C., D’Lauro, C., Clugston, J.R., McAllister, T.W., McCrea, M., Broglio, S.P.: Medical disqualification following concussion in collegiate student-athletes: findings from the care consortium. Sports medicine 50, 1843–1855 (2020)

[46] Buckley, T.A., Munkasy, B.A., Clouse, B.P.: Sensitivity and specificity of the modified balance error scoring system in concussed collegiate student athletes. Clinical journal of sport medicine 28(2), 174–176 (2018)

[47] Carlson, C.D., Langdon, J.L., Munkasy, B.A., Evans, K.M., Buckley, T.A.: Minimal detectable change scores and reliability of the balance error scoring system in student-athletes with acute concussion. Athletic Training & Sports Health Care 12(2), 67–73 (2020)

[48] Burk, J.M., Munkasy, B.A., Joyner, A.B., Buckley, T.A.: Balance error scoring system performance changes after a competitive athletic season. Clinical journal of sport medicine 23(4), 312–317 (2013)

[49] Oldham, J.R., Difabio, M.S., Kaminski, T.W., Dewolf, R.M., Howell, D.R., Buckley, T.A.: E”cacy of tandem gait to identify impaired postural control after concussion. Medicine and science in sports and exercise 50(6), 1162–1168 (2018)

[50] Howell, D.R., Buckley, T.A., Lynall, R.C., Meehan III, W.P.: Worsening dual-task gait costs after concussion and their association with subsequent sport-related injury. Journal of neurotrauma 35(14), 1630–1636 (2018)

[51] Oldham, J.R., DiFabio, M.S., Kaminski, T.W., DeWolf, R.M., Buckley, T.A.: Normative tandem gait in collegiate student-athletes: implications for clinical concussion assessment. Sports health 9(4), 305–311 (2017)

[52] Howell, D.R., Oldham, J.R., Meehan III, W.P., DiFabio, M.S., Buckley, T.A.: Dual-task tandem gait and average walking speed in healthy collegiate athletes. Clinical Journal of Sport Medicine 29(3), 238–244 (2019)

[53] Czerniak, L.L., Liebel, S.W., Garcia, G.-G.P., Lavieri, M.S., McCrea, M.A., McAllister, T.W., Broglio, S.P.: Sensitivity and specificity of computer-based neurocognitive tests in sport-related concussion: findings from the ncaa-dod care consortium. Sports medicine 51, 351–365 (2021)

[54] Czerniak, L.L., Liebel, S.W., Zhou, H., Garcia, G.-G.P., Lavieri, M.S., McCrea, M.A., McAllister, T.W., Pasquina, P.F., Broglio, S.P.: Sensitivity and specificity of the impact neurocognitive test in collegiate athletes and us military service academy cadets with adhd and/or ld: Findings from the ncaa-dod care consortium. Sports Medicine 53(3), 747–759 (2023)

[55] Tombaugh, T.N.: Trail making test a and b: normative data stratified by age and education. Archives of clinical neuropsychology 19(2), 203–214 (2004)

[56] Whitney, S.L., Eagle, S.R., Marchetti, G., Mucha, A., Collins, M.W., Kontos, A.P., Investigators, C.C.: Association of acute vestibular/ocular motor screening scores to prolonged recovery in collegiate athletes following sport-related concussion. Brain injury 34(6), 842–847 (2020)

[57] Breedlove, K.M., Ortega, J.D., Kaminski, T.W., Harmon, K.G., Schmidt, J.D., Kontos, A.P., Clugston, J.R., Chrisman, S.P., McCrea, M.A., McAllister, T.W., et al.: King-devick test reliability in national collegiate athletic association athletes: a national collegiate athletic association–department of defense concussion assessment, research and education report. Journal of athletic training 54(12), 1241–1246 (2019)

[58] Clugston, J.R., Houck, Z.M., Asken, B.M., Boone, J.K., Kontos, A.P., Buckley, T.A., Schmidt, J.D., Chrisman, S.P., Hoffman, N.L., Harmon, K.G., et al.: Relationship between the king-devick test and commonly used concussion tests at baseline. Journal of athletic training 54(12), 1247–1253 (2019)

[59] Eagle, S.R., Ferris, L.M., Mucha, A., Sinnott, A., Marchetti, G., Trbovich, A., Port, N., Clugston, J., Ortega, J., Collins, M.W., et al.: Minimum detectable change and false positive rates of the vestibular/ocular motor screening (voms) tool: an ncaa-dod care consortium analysis. Brain injury 35(12-13), 1563–1568 (2021)

[60] Caccese, J.B., Eckner, J.T., Franco-MacKendrick, L., Hazzard, J.B., Ni, M., Broglio, S.P., McAllister, T.W., McCrea, M., Buckley, T.A.: Clinical reaction-time performance factors in healthy collegiate athletes. Journal of athletic training 55(6), 601–607 (2020)

[61] Caccese, J.B., Eckner, J.T., Franco-MacKendrick, L., Hazzard, J.B., Ni, M., Broglio, S.P., McAllister, T.W., McCrea, M.A., Pasquina, P.F., Buckley, T.A.: Interpreting clinical reaction time change and recovery after concussion: a baseline versus norm-based cutoff score comparison. Journal of athletic training 56(8), 851–859 (2021)

[62] McAllister, T.W., Kenny, R., Harezlak, J., Harland, J., McCrea, M.A., Pasquina, P., Broglio, S.P., Investigators, C.C.: Profile of brief symptom inventory-18 (bsi-18) scores in collegiate athletes: A care consortium study. The Clinical Neuropsychologist, 1–16 (2024)

[63] Weber, M., Lynall, R., Hoffman, N., Miller, E., Kaminski, T., Buckley, T., Benjamin, H., Miles, C., Whitlow, C., Lintner, L., et al.: Health-related quality of life following concussion in collegiate student-athletes with and without concussion history. Annals of biomedical engineering 47, 2136–2146 (2019)

[64] Weber, M.L., Dean, J.-H.L., Hoffman, N.L., Broglio, S.P., McCrea, M., McAllister, T.W., Schmidt, J.D., Investigators, C.C., Hoy, A.R., Hazzard, J.B., et al.: Influences of mental illness, current psychological state, and concussion history on baseline concussion assessment performance. The American journal of sports medicine 46(7), 1742–1751 (2018)

[65] Anderson, M., Claros, C.C., Qian, W., Brockmeier, A., Buckley, T.A.: Integrative data analysis to identify persistent post-concussion deficits and subsequent musculoskeletal injury risk: project structure and methods. BMJ Open Sport & Exercise Medicine 10(1), 001859 (2024)

[66] Zaidi, N.A., Cerquides, J., Carman, M.J., Webb, G.I.: Alleviating naive bayes attribute independence assumption by attribute weighting (2013)

[67] Burnham, K.P., Anderson, D.R.: Model Selection and Multimodel Inference: a Practical Information-theoretic Approach. Springer, Heidelberg, Germany (2002)

[68] Zou, H., Hastie, T., Tibshirani, R.: On the “degrees of freedom” of the lasso (2007)

[69] Zhang, Y., Li, R., Tsai, C.-L.: Regularization parameter selections via generalized information criterion. Journal of the American statistical Association 105(489), 312–323 (2010)

[70] Howell, D.R., Seehusen, C.N., Carry, P.M., Walker, G.A., Reinking, S.E., Wilson, J.C.: An 8-week neuromuscular training program after concussion reduces 1-year subsequent injury risk: a randomized clinical trial. The American Journal of Sports Medicine 50(4), 1120–1129 (2022)

[71] Webster, K.E., Hewett, T.E.: Meta-analysis of meta-analyses of anterior cruciate ligament injury reduction training programs. Journal of Orthopaedic Research® 36(10), 2696–2708 (2018)

[72] Buckley, T.A., Burdette, G., Kelly, K.: Concussion-management practice patterns of national collegiate athletic association division ii and iii athletic trainers: how the other half lives. Journal of athletic training 50(8), 879–888 (2015)

[73] Kelly, K.C., Jordan, E.M., Joyner, A.B., Burdette, G.T., Buckley, T.A.: National collegiate athletic association division i athletic trainers’ concussion-management practice patterns. Journal of athletic training 49(5), 665–673 (2014)

[74] Slocum, C., Langdon, J.L., Munkasy, B.A., Brewer, B., Oldham, J.R., Graham, V., Buckley, T.A.: Multifaceted concussion assessment battery: sensitivity at the expense of specificity? The Physician and Sportsmedicine (just-accepted) (2024)

[75] Broglio, S.P., Katz, B.P., Zhao, S., McCrea, M., McAllister, T.: Test-retest reliability and interpretation of common concussion assessment tools: findings from the ncaa-dod care consortium. Sports Medicine 48, 1255–1268 (2018)

[76] Pandey, H.S., Lahijanian, B., Schmidt, J.D., Lynall, R.C., Broglio, S.P., McAllister, T.W., McCrea, M.A., Pasquina, P.F., Garcia, G.-G.P., Investigators, C.C.: Quantifying the diagnostic utility of baseline testing in concussion management: An analysis of collegiate athletes from the ncaa-dod care consortium dataset. The American Journal of Sports Medicine 53(1), 181–191 (2025)

[77] Buckley, T.A., Bryk, K.N., Hunzinger, K.J., Costantini, K.: National collegiate athletic association athletic trainers’ response to the arrington settlement: management, compliance, and practice patterns. The Physician and Sportsmedicine 51(5), 427–433 (2023)

[78] Zemek, R., Barrowman, N., Freedman, S.B., Gravel, J., Gagnon, I., McGahern, C., Aglipay, M., Sangha, G., Boutis, K., Beer, D., et al.: Clinical risk score for persistent postconcussion symptoms among children with acute concussion in the ed. Jama 315(10), 1014–1025 (2016)

[79] Refaat, M.: Credit Risk Scorecard: Development and Implementation Using SAS. Lulu, Morrisville, North Carolina (2011)

[80] Kerber, R.: Chimerge: Discretization of numeric attributes. In: Proceedings of the Tenth National Conference on Artificial Intelligence, pp. 123–128 (1992)

[81] Navas-Palencia, G.: Optimal binning: mathematical programming formulation. arXiv preprint arXiv:2001.08025 (2020)

[82] Hoerl, A.E., Kennard, R.W.: Ridge regression: applications to nonorthogonal problems. Technometrics 12(1), 69–82 (1970)

[83] Tibshirani, R.: Regression shrinkage and selection via the lasso. Journal of the Royal Statistical Society Series B: Statistical Methodology 58(1), 267–288 (1996)

[84] Sugiura, N.: Further analysis of the data by akaike’s information criterion and the finite corrections: further analysis of the data by akaike’s. Communications in Statistics-theory and Methods 7(1), 13–26 (1978)

[85] Hurvich, C.M., Tsai, C.-L.: Regression and time series model selection in small samples. Biometrika 76(2), 297–307 (1989)

[86] McCrory, P., Johnston, K., Meeuwisse, W., Aubry, M., Cantu, R., Dvorak, J., Graf-Baumann, T., Kelly, J., Lovell, M., Schamasch, P.: Summary and agreement statement of the 2nd international conference on concussion in sport, prague 2004. Clinical Journal of Sport Medicine 15(2), 48–55 (2005)

