## Supplemental material for "A Machine Learning Model for Post-Concussion Musculoskeletal Injury Risk in Collegiate Athletes"

### Appendix A Detailed Methods

This section describes the details to the statistical modeling methodology. Let $Y \in \{0,1\}$ be the binary random variable indicating subsequent injury and $\boldsymbol{x} = [x_1, \ldots, x_P] \in \mathcal{X}_1 \times \cdots \times \mathcal{X}_P$ be the vector of $P$ variables. $\boldsymbol{x}, Y \sim \mathbb{P}(\boldsymbol{x}, Y)$ are jointly distributed. The data set consists of a sample of size $N$ is $\{(y_n, \boldsymbol{x}_n)\}_{n=1}^N$, with $N = N_0 + N_1$, $N_0 = |\{n \in \{1,\ldots,N\} \ : \ y_n = 0\}|$, and $N_1 = |\{n \in \{1,\ldots,N\} \ : \ y_n = 1\}|$.

#### A.1 Naive Bayes Classifier, Weight of Evidence, and Logistic Regression

This section motivates the use of the WoE transformation by describing how it arises from a Bayesian perspective, and its similarities with the formulation of a logistic regression model.

By Bayes rule, the log-odds in favor of $Y = 1$ given variables $\boldsymbol{x} \in \mathcal{A} \subseteq \mathcal{X}_1 \times \cdots \times \mathcal{X}_P$, i.e., the variables $\boldsymbol{x}$ are in a set $\mathcal{A}$, is defined by

$$\log \frac{\mathbb{P}(Y=1|\boldsymbol{x})}{\mathbb{P}(Y=0|\boldsymbol{x})} = \log \frac{\mathbb{P}(Y=1)\mathbb{P}(\boldsymbol{x} \in \mathcal{A}|Y=1)}{\mathbb{P}(Y=0)\mathbb{P}(\boldsymbol{x} \in \mathcal{A}|Y=0)}. \tag{A1}$$

Under the naive Bayes independence assumption we assume the set is a Cartesian product $\mathcal{A} = \mathcal{B}^1 \times \cdots \times \mathcal{B}^P$ and $\mathbb{P}(\boldsymbol{x} \in \mathcal{A}|Y) = \prod_{i=1}^P \mathbb{P}(x_i \in \mathcal{A}_i|Y)$. The log-odds of $Y = 1$ yields the naive Bayes classifier's decision value

$$\log \frac{\mathbb{P}(Y=1|\boldsymbol{x})}{\mathbb{P}(Y=0|\boldsymbol{x})} = \log \frac{\mathbb{P}(Y=1)}{\mathbb{P}(Y=0)} + \sum_i \log \frac{\mathbb{P}(x_i \in \mathcal{A}_i|Y=1)}{\mathbb{P}(x_i \in \mathcal{A}_i|Y=0)}. \tag{A2}$$

The log-ratio $g_i(x_i \in \mathcal{A}_i) = \log \frac{\mathbb{P}(x_i \in \mathcal{A}_i|Y=1)}{\mathbb{P}(x_i \in \mathcal{A}_i|Y=0)}$ is called the "naive effect" (since it ignores the dependence between the $x_i$ and other variables).

In practice, the probability distributions are unknown and must be estimated from data. When the variable $x_i$ is originally continuous $\mathcal{X}_i \subseteq \mathbb{R}$, it is first binned into $C_i$ non-overlapping bins and the index of the bin is treated as a discrete random variable (see Section A.2). When $x_i$ is a discrete random variable taking $C_i$ possible values, we assume without loss of generality the values form the set $\mathcal{X}_i = \{1, \ldots, C_i\}$, the conditional distributions $\mathbb{P}(x_i = B|Y=1)$, $\mathbb{P}(x_i = B|Y=0)$, $B \in \mathcal{X}_i$, are estimated from counts $\hat{p}_{x_i|1}(B) = \frac{N_{i|1}(B)}{N_1}$ and $\hat{p}_{x_i|0}(B) = \frac{N_{i|0}(B)}{N_0}$, where $N_{i|y}(B)$ is the number of times variable $x_i = B$ when $Y = y$ and $N_y$ is the number of times $Y = y$ in the data set of size $N = N_1 + N_0$. The estimates of the naive effects using the plug-in estimates are popularly known as the *Weight of Evidence (WoE)*,

$$WoE_i(x) = \hat{g}_i(x) = \log \frac{\hat{p}_{x_i|1}(x)}{\hat{p}_{x_i|0}(x)}. \tag{A3}$$

Under the naive Bayes independence assumption, the log-odds can be expressed as

$$\log \frac{\mathbb{P}(Y=1|\boldsymbol{x})}{\mathbb{P}(Y=0|\boldsymbol{x})} = w_0 + \sum_{i=1}^{P} WoE_i(x_i), \tag{A4}$$

where $w_0 = \log \frac{\mathbb{P}(Y=1)}{\mathbb{P}(Y=0)}$ can be estimated or set using prior knowledge.

In contrast, the logistic regression model for the log-odds ratio is

$$\log \frac{\mathbb{P}(Y=1|\boldsymbol{x})}{\mathbb{P}(Y=0|\boldsymbol{x})} \approx w_0 + \sum_{i=1}^{P} w_i x_i, \tag{A5}$$

where $w_0$ is the intercept and $\sum_{i=1}^{P} w_i x_i$ is a linear combination of the variables $x_i$ with coefficients $[w_i]_{i=1}^{P}$.

For this work, we combine a logistic regression model with the WoE transformation. (The comprehensive survey [66] discusses more general approach for reweighting each variable and value, along with methods for variable selection.)

$$\log \frac{\mathbb{P}(Y=1|\boldsymbol{x})}{\mathbb{P}(Y=0|\boldsymbol{x})} \approx w_0 + \sum_{i=1}^{P} w_i \, WoE_i(x_i). \tag{A6}$$

This model can be efficiently estimated by first computing the WoE transformation on each variable, and then fitting a logistic regression model on the transformed variables, which yields an estimate of $\mathbb{P}(Y=y|\boldsymbol{x})$ given by

$$\begin{aligned} \mathbb{P}(Y=1|\boldsymbol{x};\boldsymbol{w}) &= \sigma\left(w_0 + \sum_i w_i \, WoE_i(x_i)\right), \\ \mathbb{P}(Y=0|\boldsymbol{x};\boldsymbol{w}) &= 1 - \mathbb{P}(Y=1|\boldsymbol{x};\boldsymbol{w}), \end{aligned} \tag{A7}$$

where $\sigma(z) = \frac{1}{1+e^{-z}}$ is the sigmoid function, and $\boldsymbol{w} = [w_0, w_1, \ldots, w_P]$ is the vector of parameters (bias and coefficients).

#### A.2 Data Preprocessing with Optimal Binning and WoE Transformation

Given a training data set of observed data points $\mathcal{D}_N = \{(y_n, \boldsymbol{x}_n)\}_{n=1}^{N}$, optimal binning strategy for each of the variables $x_i$ in the data set involves finding the best splits that maximize the Information Value (IV). For categorical variables, maximizing the IV may require regrouping the categories. For ordinal and continuous variables, maximizing the IV depends on creating split points. Once the bins that optimize the IV are defined, we apply the WoE transformation and replace the original variable values with the log-ratio value at each bin, converting the variables into more informative representations suitable for predictive modeling.

Since the WoE transformation is defined for discrete variables, we need to find an appropriate representation for continuous variables that can be used in this framework. When the variable $x_i$ is originally continuous $\mathcal{X}_i \subseteq \mathbb{R}$, it is first binned into $C_i$ non-overlapping bins, $\mathcal{B}^i_1, \ldots, \mathcal{B}^i_{C_i}$, $\bigcup_j \mathcal{B}^i_j = \mathbb{R}$, $j \neq k \implies \mathcal{B}^i_j \cap \mathcal{B}^i_k = \emptyset$, with the binning function $\mathfrak{b}_i(x) = j \iff x \in \mathcal{B}^i_j$. Then, $\mathfrak{b}_i(x_i) \in \{1, \ldots, C_i\}$ is a discrete random variable and the conditional distributions $\mathbb{P}(x_i \in \mathcal{B}^i_{\mathfrak{b}_i(x)} | Y = y), \quad x \in \mathcal{X}_i \subseteq \mathbb{R}, y \in \{0, 1\}$ are estimated by counts $\hat{p}_{x_i|y}(x) = \frac{N_{i|y}(\mathfrak{b}_i(x))}{N_y}$. After binning, the WoE transformation for variable $x_i$ is a piecewise constant function.

Explicitly, the $C_i$ bins, and the binning function $\mathfrak{b}_i : \mathbb{R} \to \{1, \ldots, C_i\}$, are defined by $C_i - 1$ split points $b^i_1, b^i_2, \ldots, b^i_{C_i-1} \in \mathbb{R}$ characterized by the intervals:

$$\Big\{ \underbrace{(-\infty, b^i_1)}_{\mathcal{B}^i_1}, \underbrace{[b^i_1, b^i_2)}_{\mathcal{B}^i_2}, \ldots, \underbrace{[b^i_{C_i-1}, +\infty)}_{\mathcal{B}^i_{C_i}} \Big\}, \text{where } b^i_1 < b^i_2 < \cdots < b^i_{C_i-1}. \tag{A8}$$

We pose the selection of the number $C_i$ and value of the split points as an optimization of the Information Value (IV) of the resulting conditional distributions, which assesses the binned variables predictive power. IV is calculated based on the WoE values of each category or bin of a predictor variable and their corresponding proportions within the classes. The formula to compute the IV for a given variable $x_i$ with $C_i$ bins is

$$\begin{aligned} IV(x_i) &= \sum_{j=1}^{C_i} \big(\hat{p}_{x_i|1}(j) - \hat{p}_{x_i|0}(j)\big) \log \frac{\hat{p}_{x_i|1}(j)}{\hat{p}_{x_i|0}(j)} \\ &= \frac{1}{N_1} \sum_{n:y_n=1} WoE_i(x_{ni}) - \frac{1}{N_0} \sum_{n:y_n=0} WoE_i(x_{ni}), \end{aligned} \tag{A9}$$

for a data set $\{(y_n, x_{ni})\}_{n=1}^{N_0+N_1}$.

IV quantifies the predictive power of a predictor variable $x_i$ by measuring the separation between the evidence in favor of the target $\hat{p}_{x_i|1}$ and the evidence against the target $\hat{p}_{x_i|0}$. Note that the term $\big(\hat{p}_{x_i|1}(j) - \hat{p}_{x_i|0}(j)\big) \log \frac{\hat{p}_{x_i|1}(j)}{\hat{p}_{x_i|0}(j)}$ is always positive. Therefore, when the IV value of a predictor variable is large, it suggests that the variable is highly predictive of the target variable. In other words, the WoE values for different categories are made to be maximally different to help distinguish the classes. Variables with high IV value are valuable for building robust predictive models. Contrastingly, when the IV value of a predictor variable is closer to 0, it implies that the variable has little discriminatory power since the evidence in favor and against each class is similar, indicating that the variable does not provide much information to differentiate between classes. Inclusion of such variables in a predictive model may not contribute significantly to its performance. In practice, the IV is often used to select the most informative predictor variables. Variables with IV values above a certain threshold are retained, while those below the threshold may be considered for further evaluation or exclusion from the model [36, 79]. For our analysis, IV is used to evaluate discriminant power of a particular binning scheme.

Finding the best representation of a variable is at the heart of this approach. In that sense, optimal binning is a data preprocessing technique that aims to divide continuous or discrete predictor variables into a set of bins or intervals in a way that maximizes the separation between the classes of the target variable. The goal is to create bins where the within-bin homogeneity (similar outcomes) and between-bin heterogeneity (different outcomes) are maximized, leading to a more predictive and informative representation of the data [42, 80].

The process of optimal binning typically involves an iterative algorithm that examines different binning configurations and evaluates their performance using a pre-defined criterion. Commonly used criteria include maximizing the IV, satisfying some constraints such as a minimum number of observations in each bin, or ensuring some monotonic behavior (increasing or decreasing trend in the target variable within the bins) in the WoE values of a variable [81].

As the $IV(x_i)$ expressed above is a function of the binning in terms of the WoE transform, defined by the split points, $b_1^i, \ldots, b_{C_i-1}^i$, the optimal binning that maximizes the IV requires a search over splits and a search over the number of splits:

$$b_1^i, \ldots, b_{C_i-1}^i = \underset{b_1, b_2, \ldots, b_{C_i}}{\arg\max}\; IV(x_i), \tag{A10}$$

$$C_i = \underset{C \in \{2, \ldots, N\}}{\arg\max}\; \max_{b_1, b_2, \ldots, b_C} IV(x_i). \tag{A11}$$

This is a combinatorial problem and an exhaustive search is not feasible. Instead, the process is divided into two steps: a pre-binning step, which generates a granular segmentation of the raw variable $x_i$, and an optimization step, which combines contiguous segments (bins) searching for a configuration that yields the highest IV, i.e, the most informative representation with respect to the target. The optimal binning and WoE transformation is carried out using the Python library OPTBINNING v0.17.3 [81].

#### A.3 Fitting the Logistic Regression Model

Given a training data set of observed data points $\mathcal{D}_N = \{(y_n \boldsymbol{x}_n)\}_{n=1}^N$, and the WoE transformations with optimal binning, $\{WoE_i\}_{i=1}^P$ model estimation of $\boldsymbol{w}$ is typically done by maximizing the likelihood function $\mathcal{L}(\boldsymbol{w}|\mathcal{D}_N) = \prod_{n=1}^N P(Y = y_n|\boldsymbol{x}_n; \boldsymbol{w})$, or equivalently, minimizing the negative log-likelihood, which in the case of logistic regression is

$$\begin{aligned} -\log(\mathcal{L}(\boldsymbol{w}|\mathcal{D}_N)) = &-\sum_{n=1}^N y_n \log(\sigma(w_0 + \sum_{i=1}^P w_i\, WoE_i(x_{n,i}))) \\ &+ (1 - y_n) \log(1 - \sigma(w_0 + \sum_{i=1}^P w_i\, WoE_i(x_{n,i}))). \end{aligned} \tag{A12}$$

The optimal values for the coefficients $\boldsymbol{w}$ are obtained as the solution to the optimization problem

$$\boldsymbol{w}^\star = \arg\min_{\boldsymbol{w}} -\log(L(\boldsymbol{w}|\mathcal{D}_N)), \tag{A13}$$

which can be efficiently performed using various solvers.

##### A.3.1 Regularized likelihood

Regularization is a technique for adding an additional penalty to the coefficients to the model fitting criterion. Considering the solution as a statistical estimate, regularized solutions have lower variance, but introduces some bias, with proper choice, regularization can improve generalization by preventing overfitting. The penalty is a function on the values of the coefficient vector: Ridge regularization [82] uses a squared L2-norm, while LASSO [83] uses an L1-norm. LASSO has the added benefit that it implicitly performs variable selection as the the $\ell_1$-norm encourages sparsity (model coefficients bin exactly zero) in the coefficient vector $\boldsymbol{w}$, leading to a more interpretable and parsimonious model. Under these considerations, the regularized logistic regression optimization problem is

$$\boldsymbol{w}^\lambda = \arg\min_{\boldsymbol{w}} -\log(L(\boldsymbol{w}|\mathcal{D}_N)) + \lambda\|\boldsymbol{w}\|_1, \tag{A14}$$

where $\|\boldsymbol{w}\|_1 = \sum_{i=1}^{P}|w_i|$ is the L1-norm and $\lambda$ is the regularization parameter. A larger value of $\lambda$ implies a stronger penalty on the coefficients, which induces variable selection. The selection of $\lambda$ is critical, and it can be performed by model selection criterion or cross-validation performance.

When the sample size is relatively small, model selection criterion are preferred. In particular if the ratio $N/P < 10$, where $N$ is the sample size and $P$ the number of variables, information criteria-based metrics can be more beneficial [67]. For small sample regimes, an appropriate choice is corrected Akaike Information Criterion (AICc)[84, 85], which is defined as

$$AICc(\theta) = -2\log L(\theta|\mathcal{D}_N) + 2K(\theta) + \frac{2K(\theta)(K(\theta)+1)}{N-K(\theta)-1} = AIC(\theta) + \frac{2K(\theta)(K(\theta)+1)}{N-K(\theta)-1}, \tag{A15}$$

where $\theta$ is a model, $L(\theta|\mathcal{D}_N)$ is the likelihood, and $K(\theta)$ is the number of parameters of the model. For a logistic regression model defined by the parameters $\boldsymbol{w}^\star$, $\theta = \boldsymbol{w}^\lambda$ and $K(\theta) = |\text{sup}(\boldsymbol{w}^\lambda)| = |\{i \in \{0, 1, \ldots, P\} : w_i^\lambda \neq 0\}|$ is the number of non-zero coefficients. Given a regularization path $\boldsymbol{w}_{\lambda_1}, \ldots \boldsymbol{w}_{\lambda_T}$, we can compute $AICc$ for a range of values $\lambda_1 < \lambda_2 < \cdots < \lambda_T$. The value of $\lambda$ that minimizes the $AICc$ balances the model fit with the number of variables used in the model,

$$\lambda^\star = \arg\min_{\lambda \in \{\lambda_t\}_{t=1}^T} \text{AICc}(\boldsymbol{w}^\lambda). \tag{A16}$$

### Appendix B Supplementary Results

To understand the WoE transformation, we visualize the WoE values across the distribution for a specific variables using a histogram.

Fig. B1 shows the behavior of the binning approach and the WoE coefficients for the variable `SCAT Severity Difference Baseline Asymptomatic`, which is a standardized tool for evaluating concussions and evaluates 22 symptoms with a 7-point Likert Scale (0=none, . . . , 6=severe) [86]. The WoE values across the bins show that the risk of injury after a concussion is greater if the athlete's SCAT score is higher at the asymptomatic time point than the SCAT score at the baseline time point. In contrast, the risk of injury post-concussion is the lowest when the athlete's asymptomatic SCAT score is lower than the baseline score.

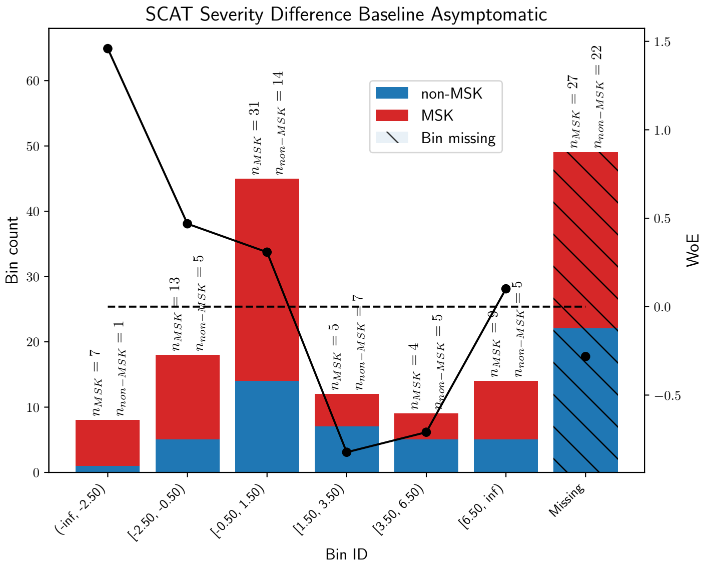


**Fig. B1**: WoE values and histograms after optimal binning of the `SCAT Severity Difference Baseline Asymptomatic` variable, which is the difference between the SCAT scores at baseline and asymptomatic time points. The leftmost bins of negative values correspond to higher SCAT scores at asymptomatic time point compared to baseline, and are associated with a higher likelihood of injury after concussion. If the SCAT score at asymptomatic time point is lower than the one at baseline, then the risk is lower or negligible. The righmost stacked bar (cross-hatched) is for the missing values, with the dot indicating a negative WoE value corresponding to decreased risk.

Fig. B2 shows the result of the feature selection process under both transformations. Table B1 depicts ranked variables by their mean absolute SHAP value calculated based on the training set (`SHAP train`). Tables B2 and B3 show an expanded version of the risk computation. Given an athlete's variables, we can use Table B2 to locate the bins that correspond to the original measurements. After finding the bins, we can find in Table B3 the associated individual score. The higher the sum of all the individual scores, the higher the risk of injury.

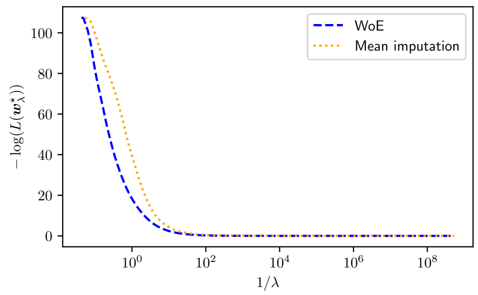


(a) Negative Log-Likelihood

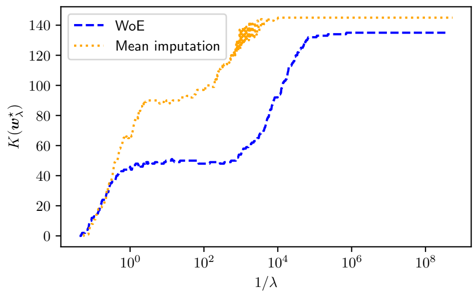


(b) Number of Non-zero Coefficients

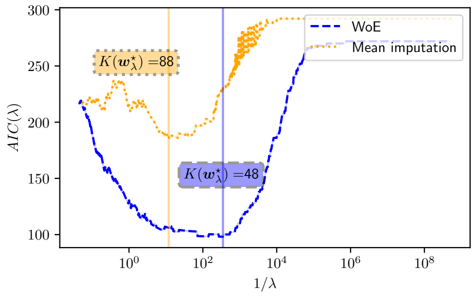


(c) AIC

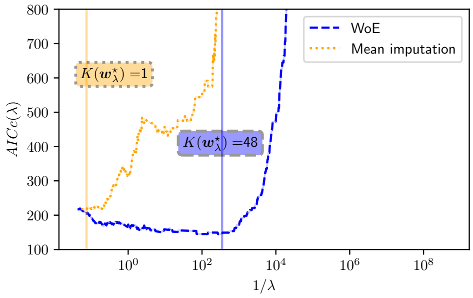


(d) AICc

**Fig. B2**: Variable selection based on L1-penalized logistic regression. Plot (a) shows that the negative log likelihood is minimized approximately for $\lambda > 10$; however the number of non-zero coefficients. Plot (b) varies widely in the same range. Plots (c) and (d) show a better picture representation of the performance in the test set. Given our sample size and number of variables, we use the AICc (Plot (d)) to pick the optimal strength for $\lambda$.

Finally, the distribution of the logit scores and the ROC and Precision-Recall curves for both our proposed approach and the benchmark model are shown in Fig. B3 and Fig. B4, respectively. Our model shows a clear superior performance compared to the benchmark model, showing that our proposed approach that encompasses WoE transformation with optimal binning followed by an information theory-based model selection procedure is able to generate a robust predictive model. The model's performance in the test set suggests that its predictive skills translates into real-world scenarios.

| $i$ | Variable $x_i$ | SHAP train | SHAP test | $w_i$ |
|---|---|---|---|---|
| 1 | TMT-A Time@Acute | 0.464 | 0.536 | 1.275 |
| 2 | BESS@Baseline | 0.413 | 0.360 | 1.186 |
| 3 | CRT Difference Baseline Acute | 0.408 | 0.401 | 0.931 |
| 4 | ImPACT Reaction Time@Asymptomatic | 0.385 | 0.277 | 0.901 |
| 5 | Sport | 0.367 | 0.299 | 0.768 |
| 6 | SWLS | 0.352 | 0.337 | 0.851 |
| 7 | HADS Depression | 0.303 | 0.255 | 0.774 |
| 8 | K-D Time@Baseline | 0.302 | 0.343 | 0.633 |
| 9 | Time Lost | 0.289 | 0.311 | 1.064 |
| 10 | K-D Time Difference Baseline Asymptomatic | 0.289 | 0.343 | 0.694 |
| 11 | ImPACT Reaction Time@RTA | 0.288 | 0.353 | 0.722 |
| 12 | NPC@Baseline | 0.282 | 0.268 | 0.723 |
| 13 | TG ST@Baseline | 0.278 | 0.280 | 0.664 |
| 14 | TMT-A Time Difference Baseline Asymptomatic | 0.259 | 0.256 | 0.955 |
| 15 | K-D Time@Acute | 0.250 | 0.339 | 0.495 |
| 16 | BSI@Acute | 0.244 | 0.240 | 0.863 |
| 17 | SCAT Severity Difference Baseline Asymptomatic | 0.233 | 0.256 | 0.720 |
| 18 | ImPACT Motor Speed@RTA | 0.228 | 0.236 | 0.572 |
| 19 | TG DT@Baseline | 0.226 | 0.201 | 0.486 |
| 20 | SAC@Asymptomatic | 0.207 | 0.238 | 0.613 |
| 21 | TMT-A Time@Baseline | 0.206 | 0.171 | 0.511 |
| 22 | TMT-B Time@Asymptomatic | 0.195 | 0.230 | 0.708 |
| 23 | NPC@Acute | 0.193 | 0.125 | 0.688 |
| 24 | ImPACT Verbal Memory Difference Baseline RTA | 0.191 | 0.195 | 0.536 |
| 25 | ImPACT Verbal Memory@RTA | 0.186 | 0.225 | 0.537 |
| 26 | K-D Time Difference Baseline RTA | 0.179 | 0.173 | 0.555 |
| 27 | TMT-B Time@Acute | 0.167 | 0.118 | 0.644 |
| 28 | TG ST@Acute | 0.150 | 0.116 | 0.487 |
| 29 | ImPACT Visual Memory Difference Baseline Acute | 0.146 | 0.184 | 0.470 |
| 30 | TG DT@RTA | 0.144 | 0.132 | 0.936 |
| 31 | K-D Time@RTA | 0.123 | 0.119 | 0.332 |
| 32 | CRT@Asymptomatic | 0.120 | 0.123 | 0.506 |
| 33 | BESS@Asymptomatic | 0.116 | 0.099 | 0.478 |
| 34 | ImPACT Reaction Time Difference Baseline Asymptomatic | 0.113 | 0.089 | 0.378 |
| 35 | TMT-B Time@Baseline | 0.112 | 0.080 | 0.323 |
| 36 | TG DT Difference Baseline RTA | 0.112 | 0.057 | 0.473 |
| 37 | ImPACT Verbal Memory Difference Baseline Acute | 0.112 | 0.104 | 0.333 |
| 38 | TG DT@Asymptomatic | 0.111 | 0.079 | 0.587 |
| 39 | ImPACT Visual Memory@Acute | 0.104 | 0.094 | 0.457 |
| 40 | ImPACT Motor Speed Difference Baseline Asymptomatic | 0.104 | 0.148 | 0.461 |
| 41 | CRT Difference Baseline RTA | 0.099 | 0.122 | 0.492 |
| 42 | ImPACT Reaction Time Difference Baseline RTA | 0.073 | 0.104 | 0.406 |
| 43 | ImPACT Motor Speed@Baseline | 0.068 | 0.061 | 0.210 |
| 44 | TMT-A Time@Asymptomatic | 0.057 | 0.098 | 0.300 |
| 45 | TMT-A Time Difference Baseline Acute | 0.056 | 0.056 | -0.265 |
| 46 | K-D Time Difference Baseline Acute | 0.036 | 0.053 | -0.168 |
| 47 | NPC@RTA | 0.026 | 0.025 | -0.177 |
| 48 | ImPACT Visual Memory@Baseline | 0.015 | 0.014 | 0.047 |

**Table B1**: Variable importance. This table shows the variables used to train the logistic regression model ranked by their absolute SHAP value based on the training set. The higher SHAP value, the more important the contribution of that variable to prediction. Additionally, the coefficients $w_i$ for each of the variables of the trained logistic regression model are displayed. The bias term value is $w_0 = -1.093$.

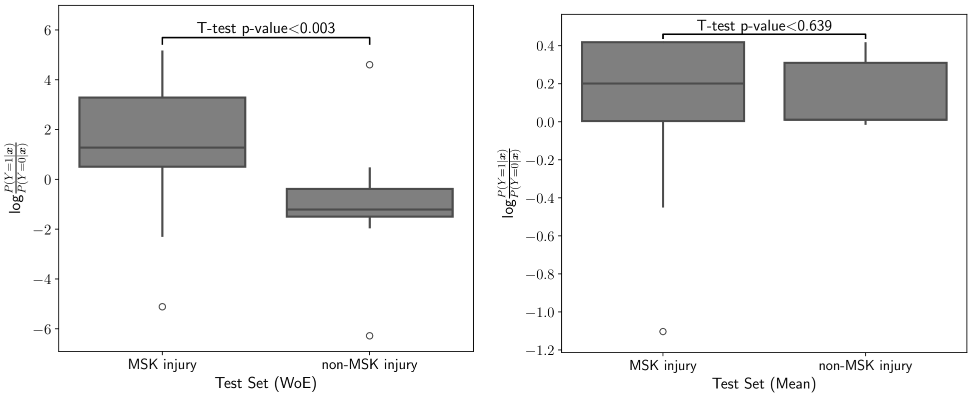


(a) Log-odds Ratio Scores for the test set when WoE transformation is used.

(b) Log-odds Ratio Scores for the test set when mean imputation is used.

**Fig. B3**: Distribution of Logit Scores for MSK injury and Non-MSK injury Groups in the Test Set. The box plot illustrate the spread and central tendency of the logit risk scores computed for our proposed approach and the benchmark model. The model trained for WoE-transformed variables (a) shows a statistical difference between the two groups, while the benchmark model (b) fails to generate statistically different predictions.

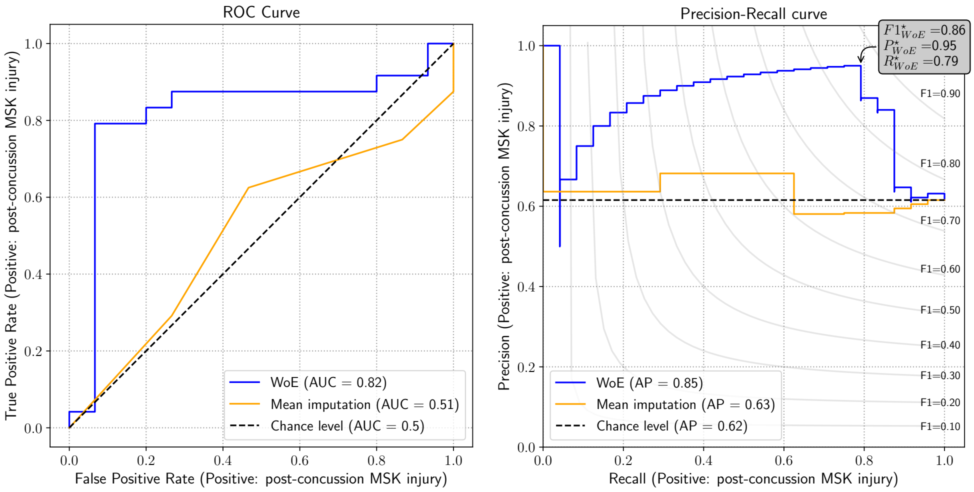


(a) Receiver Operating Characteristic Curve

(b) Precision-Recall Curve.

**Fig. B4**: The ROC and Precision-Recall Curves for the Logistic Regression Models Post Variable Selection for our proposed model and the benchmark. Our model trained using WoE-transformed variables displays a stronger performance in contrast to the benchmark model.

| $i$ | Variable | $\mathcal{B}_1^i$ | $\mathcal{B}_2^i$ | $\mathcal{B}_3^i$ | $\mathcal{B}_4^i$ | $\mathcal{B}_5^i$ | $\mathcal{B}_6^i$ | $\mathcal{B}_7^i$ | $\mathcal{B}_8^i$ | $\mathcal{B}_9^i$ | $\mathcal{B}_{10}^i$ |
|---|---|---|---|---|---|---|---|---|---|---|---|
| 1 | BESS@Asymptomatic | (-inf, 4.50) | [4.50, 6.50) | [6.50, 11.50) | [11.50, 14.50) | [14.50, inf) | Missing | | | | |
| 2 | BESS@Baseline | (-inf, 5.50) | [5.50, 7.50) | [7.50, 8.50) | [8.50, 11.50) | [11.50, 20.50) | [20.50, inf) | Missing | | | |
| 3 | BSI@Acute | (-inf, 5.50) | [5.50, 8.50) | [8.50, inf) | Missing | | | | | | |
| 4 | CRT Difference Baseline Acute | (-inf, -50.50) | [-50.50, -32.50) | [-32.50, -12.50) | [-12.50, 15.50) | [15.50, inf) | Missing | | | | |
| 5 | CRT Difference Baseline RTA | (-inf, -19.50) | [-19.50, 38.50) | [38.50, inf) | Missing | | | | | | |
| 6 | CRT@Asymptomatic | (-inf, 137.50) | [137.50, 151.50) | [151.50, 154.50) | [154.50, 189.00) | [189.00, inf) | Missing | | | | |
| 7 | HADS Depression | (-inf, 0.50) | [0.50, 1.50) | [1.50, 2.50) | [2.50, 6.50) | [6.50, inf) | Missing | | | | |
| 8 | ImPACT Motor Speed Difference Baseline Asymptomatic | (-inf, -7.55) | [-7.55, -0.12) | [-0.12, 2.88) | [2.88, 4.78) | [4.78, inf) | Missing | | | | |
| 9 | ImPACT Motor Speed@Baseline | (-inf, 34.85) | [34.85, 37.12) | [37.12, 49.27) | [49.27, inf) | Missing | | | | | |
| 10 | ImPACT Motor Speed@RTA | (-inf, 35.23) | [35.23, 37.49) | [37.49, 42.56) | [42.56, 46.05) | [46.05, inf) | Missing | | | | |
| 11 | ImPACT Reaction Time Difference Baseline Asymptomatic | (-inf, -0.11) | [-0.11, -0.02) | [-0.02, -0.01) | [-0.01, 0.01) | [0.01, 0.06) | [0.06, 0.11) | [0.11, inf) | Missing | | |
| 12 | ImPACT Reaction Time Difference Baseline RTA | (-inf, -0.09) | [-0.09, -0.03) | [-0.03, -0.01) | [-0.01, inf) | Missing | | | | | |
| 13 | ImPACT Reaction Time@Asymptomatic | (-inf, 0.51) | [0.51, 0.56) | [0.56, 0.57) | [0.57, 0.64) | [0.64, inf) | Missing | | | | |
| 14 | ImPACT Reaction Time@RTA | (-inf, 0.50) | [0.50, 0.56) | [0.56, 0.63) | [0.63, 0.69) | [0.69, inf) | Missing | | | | |
| 15 | ImPACT Verbal Memory Difference Baseline Acute | (-inf, -12.00) | [-12.00, -3.50) | [-3.50, 0.50) | [0.50, 3.50) | [3.50, 9.50) | [9.50, inf) | Missing | | | |
| 16 | ImPACT Verbal Memory Difference Baseline RTA | (-inf, -16.50) | [-16.50, -12.50) | [-12.50, -9.00) | [-9.00, 7.00) | [7.00, inf) | Missing | | | | |
| 17 | ImPACT Verbal Memory@RTA | (-inf, 74.00) | [74.00, 86.50) | [86.50, 91.50) | [91.50, 93.50) | [93.50, inf) | Missing | | | | |
| 18 | ImPACT Visual Memory Difference Baseline Acute | (-inf, -12.50) | [-12.50, -6.50) | [-6.50, 21.50) | [21.50, inf) | Missing | | | | | |
| 19 | ImPACT Visual Memory@Acute | (-inf, 51.50) | [51.50, 60.50) | [60.50, 67.50) | [67.50, 89.50) | [89.50, inf) | Missing | | | | |
| 20 | ImPACT Visual Memory@Baseline | (-inf, 72.50) | [72.50, 74.50) | [74.50, 76.50) | [76.50, 78.50) | [78.50, 81.50) | [81.50, inf) | Missing | | | |
| 21 | K-D Time Difference Baseline Acute | (-inf, -11.45) | [-11.45, inf) | Missing | | | | | | | |
| 22 | K-D Time Difference Baseline Asymptomatic | (-inf, -4.59) | [-4.59, 3.55) | [3.55, 5.42) | [5.42, 8.32) | [8.32, 10.46) | [10.46, inf) | Missing | | | |
| 23 | K-D Time Difference Baseline RTA | (-inf, 1.68) | [1.68, 3.47) | [3.47, inf) | Missing | | | | | | |
| 24 | K-D Time@Acute | (-inf, 35.08) | [35.08, 53.07) | [53.07, 61.95) | [61.95, inf) | Missing | | | | | |
| 25 | K-D Time@Baseline | (-inf, 34.87) | [34.87, 48.44) | [48.44, 52.50) | [52.50, inf) | Missing | | | | | |
| 26 | K-D Time@RTA | (-inf, 30.57) | [30.57, 35.90) | [35.90, 37.06) | [37.06, 42.22) | [42.22, 44.59) | [44.59, inf) | Missing | | | |
| 27 | NPC@Acute | (-inf, 5.33) | [5.33, 7.25) | [7.25, inf) | Missing | | | | | | |
| 28 | NPC@Baseline | (-inf, 1.75) | [1.75, 2.54) | [2.54, 4.83) | [4.83, inf) | Missing | | | | | |
| 29 | NPC@RTA | (-inf, 1.17) | [1.17, 2.33) | [2.33, inf) | Missing | | | | | | |
| 30 | SAC@Asymptomatic | (-inf, 25.50) | [25.50, 26.50) | [26.50, 27.50) | [27.50, 28.50) | [28.50, inf) | Missing | | | | |
| 31 | SCAT Severity Difference Baseline Asymptomatic | (-inf, -2.50) | [-2.50, -0.50) | [-0.50, 1.50) | [1.50, 3.50) | [3.50, 6.50) | [6.50, inf) | Missing | | | |
| 32 | SWLS | (-inf, 19.50) | [19.50, 25.50) | [25.50, 26.50) | [26.50, 28.50) | [28.50, 29.50) | [29.50, inf) | Missing | | | |
| 33 | Sport | ['Swimming and Diving'] | ['Rowing'] | ['Tennis'] ['Lacrosse'] ['Cheer'] | ['Football'] | ['Soccer'] | ['Baseball and Softball'] | ['Basketball'] | ['Volleyball'] | ['Track and Field'] | ['Field Hockey'] |
| 34 | TG DT Difference Baseline RTA | (-inf, 0.47) | [0.47, 4.03) | [4.03, inf) | Missing | | | | | | |
| 35 | TG DT@Asymptomatic | (-inf, 10.67) | [10.67, 11.31) | [11.31, inf) | Missing | | | | | | |
| 36 | TG DT@Baseline | (-inf, 9.48) | [9.48, 11.79) | [11.79, 12.68) | [12.68, 13.88) | [13.88, 19.10) | [19.10, inf) | Missing | | | |
| 37 | TG DT@RTA | (-inf, 10.79) | [10.79, 13.03) | [13.03, 14.79) | [14.79, inf) | Missing | | | | | |
| 38 | TG ST@Acute | (-inf, 8.53) | [8.53, 10.23) | [10.23, 10.85) | [10.85, inf) | Missing | | | | | |
| 39 | TG ST@Baseline | (-inf, 10.53) | [10.53, 11.68) | [11.68, 12.69) | [12.69, inf) | Missing | | | | | |
| 40 | TMT-A Time Difference Baseline Acute | (-inf, -5.25) | [-5.25, 3.25) | [3.25, 6.85) | [6.85, inf) | Missing | | | | | |
| 41 | TMT-A Time Difference Baseline Asymptomatic | (-inf, 0.85) | [0.85, 2.05) | [2.05, 3.10) | [3.10, 6.85) | [6.85, inf) | Missing | | | | |
| 42 | TMT-A Time@Acute | (-inf, 15.20) | [15.20, 15.90) | [15.90, 17.85) | [17.85, 19.50) | [19.50, 23.00) | [23.00, inf) | Missing | | | |
| 43 | TMT-A Time@Asymptomatic | (-inf, 13.85) | [13.85, 21.45) | [21.45, inf) | Missing | | | | | | |
| 44 | TMT-A Time@Baseline | (-inf, 14.35) | [14.35, 15.45) | [15.45, 18.45) | [18.45, 19.75) | [19.75, 21.85) | [21.85, inf) | Missing | | | |
| 45 | TMT-B Time@Acute | (-inf, 29.70) | [29.70, 41.55) | [41.55, 43.10) | [43.10, 45.30) | [45.30, 52.50) | [52.50, inf) | Missing | | | |
| 46 | TMT-B Time@Asymptomatic | (-inf, 29.60) | [29.60, 32.05) | [32.05, 35.05) | [35.05, 45.45) | [45.45, inf) | Missing | | | | |
| 47 | TMT-B Time@Baseline | (-inf, 31.20) | [31.20, 37.05) | [37.05, 38.95) | [38.95, 54.65) | [54.65, inf) | Missing | | | | |
| 48 | Time Lost | (-inf, 9.50) | [9.50, 10.50) | [10.50, 11.50) | [11.50, inf) | | | | | | |

**Table B2**: Optimal binning. This table shows the resulting binning configuration for each of the variables that were picked after the feature selection process and used to train the logistic regression model. Variables are arranged alphabetically.

| $i$ | Variable | $w_i\mathcal{B}^i_1$ | $w_i\mathcal{B}^i_2$ | $w_i\mathcal{B}^i_3$ | $w_i\mathcal{B}^i_4$ | $w_i\mathcal{B}^i_5$ | $w_i\mathcal{B}^i_6$ | $w_i\mathcal{B}^i_7$ | $w_i\mathcal{B}^i_8$ | $w_i\mathcal{B}^i_9$ | $w_i\mathcal{B}^i_{10}$ |
|---|---|---|---|---|---|---|---|---|---|---|---|
| 1 | BESS@Asymptomatic | -0.126 | -0.427 | 0.035 | 0.132 | 0.216 | -0.087 | | | | |
| 2 | BESS@Baseline | 0.726 | -0.236 | -0.313 | -0.955 | 0.335 | 0.802 | -0.577 | | | |
| 3 | BSI@Acute | 0.267 | -0.710 | 1.051 | -0.358 | | | | | | |
| 4 | CRT Difference Baseline Acute | 0.336 | -1.476 | 0.267 | 0.373 | 1.592 | -0.352 | | | | |
| 5 | CRT Difference Baseline RTA | -0.381 | 0.102 | 0.301 | -0.052 | | | | | | |
| 6 | CRT@Asymptomatic | 0.153 | -0.179 | -0.359 | 0.058 | 0.373 | -0.133 | | | | |
| 7 | HADS Depression | 0.629 | 0.007 | -0.063 | -0.174 | -1.033 | -0.236 | | | | |
| 8 | ImPACT Motor Speed Difference Baseline Asymptomatic | 0.282 | 0.095 | -0.016 | -0.482 | 0.353 | -0.064 | | | | |
| 9 | ImPACT Motor Speed@Baseline | -0.004 | -0.393 | 0.040 | 0.291 | -0.149 | | | | | |
| 10 | ImPACT Motor Speed@RTA | -0.510 | -0.907 | -0.321 | 0.014 | 0.262 | 0.234 | | | | |
| 11 | ImPACT Reaction Time Difference Baseline Asymptomatic | 0.009 | -0.085 | -0.377 | 0.009 | 0.108 | 0.198 | 0.552 | -0.052 | | |
| 12 | ImPACT Reaction Time Difference Baseline RTA | 0.311 | -0.479 | -0.198 | -0.040 | 0.076 | | | | | |
| 13 | ImPACT Reaction Time@Asymptomatic | 1.722 | 0.341 | -1.063 | -0.387 | 0.022 | 0.022 | | | | |
| 14 | ImPACT Reaction Time@RTA | 0.553 | 0.028 | -0.496 | -1.221 | 0.149 | 0.295 | | | | |
| 15 | ImPACT Verbal Memory Difference Baseline Acute | -0.213 | 0.017 | 0.092 | 0.605 | 0.181 | 0.034 | -0.127 | | | |
| 16 | ImPACT Verbal Memory Difference Baseline RTA | -1.304 | 0.193 | 0.782 | -0.033 | -0.381 | 0.100 | | | | |
| 17 | ImPACT Verbal Memory@RTA | -1.306 | -0.396 | 0.329 | 0.111 | -0.076 | 0.220 | | | | |
| 18 | ImPACT Visual Memory Difference Baseline Acute | 0.288 | -0.745 | 0.108 | 0.651 | -0.179 | | | | | |
| 19 | ImPACT Visual Memory@Acute | 0.011 | -0.408 | 0.055 | 0.169 | 0.178 | -0.183 | | | | |
| 20 | ImPACT Visual Memory@Baseline | 0.006 | -0.023 | -0.055 | -0.033 | 0.017 | 0.022 | -0.033 | | | |
| 21 | K-D Time Difference Baseline Acute | 0.168 | -0.055 | 0.035 | | | | | | | |
| 22 | K-D Time Difference Baseline Asymptomatic | 0.845 | 0.199 | -1.300 | -0.183 | 0.425 | 0.532 | -0.124 | | | |
| 23 | K-D Time Difference Baseline RTA | 0.479 | -0.880 | -0.025 | -0.086 | | | | | | |
| 24 | K-D Time@Acute | -0.442 | 0.197 | 1.065 | -0.785 | -0.189 | | | | | |
| 25 | K-D Time@Baseline | -0.193 | 0.176 | 1.265 | -1.131 | -0.229 | | | | | |
| 26 | K-D Time@RTA | -0.392 | -0.127 | 0.568 | 0.150 | 0.069 | -0.027 | -0.043 | | | |
| 27 | NPC@Acute | 0.120 | -0.918 | 0.661 | -0.100 | | | | | | |
| 28 | NPC@Baseline | 0.011 | 0.442 | 1.502 | -0.645 | -0.164 | | | | | |
| 29 | NPC@RTA | -0.013 | 0.169 | -0.013 | -0.019 | | | | | | |
| 30 | SAC@Asymptomatic | 1.113 | 0.287 | 0.081 | -0.256 | -0.050 | -0.144 | | | | |
| 31 | SCAT Severity Difference Baseline Asymptomatic | 1.051 | 0.337 | 0.222 | -0.593 | -0.511 | 0.073 | -0.203 | | | |
| 32 | SWLS | -1.135 | 0.051 | 0.062 | 1.890 | 0.176 | 0.044 | -0.473 | | | |
| 33 | Sport | -0.906 | -0.766 | -0.288 | -0.001 | 0.018 | 0.056 | 0.330 | 0.470 | 0.781 | 1.223 |
| 34 | TG DT Difference Baseline RTA | -0.422 | 0.617 | -0.316 | 0.011 | | | | | | |
| 35 | TG DT@Asymptomatic | -0.524 | 0.856 | 0.070 | 0.008 | | | | | | |
| 36 | TG DT@Baseline | -0.128 | 0.358 | 0.512 | 0.494 | -0.172 | -0.485 | -0.143 | | | |
| 37 | TG DT@RTA | 0.337 | -0.835 | 0.022 | 0.462 | -0.024 | | | | | |
| 38 | TG ST@Acute | 0.438 | -0.162 | -0.772 | 0.247 | -0.044 | | | | | |
| 39 | TG ST@Baseline | 0.376 | -0.011 | -0.886 | 1.058 | -0.323 | | | | | |
| 40 | TMT-A Time Difference Baseline Acute | -0.162 | -0.120 | -0.019 | 0.218 | 0.024 | | | | | |
| 41 | TMT-A Time Difference Baseline Asymptomatic | 0.859 | -0.465 | -1.514 | 0.023 | 0.197 | -0.028 | | | | |
| 42 | TMT-A Time@Acute | 0.496 | -1.272 | -1.050 | 0.031 | 0.548 | 1.147 | -0.211 | | | |
| 43 | TMT-A Time@Asymptomatic | 0.097 | 0.040 | -0.268 | -0.010 | | | | | | |
| 44 | TMT-A Time@Baseline | 0.746 | 0.313 | 0.193 | -0.864 | -0.018 | 0.191 | -0.156 | | | |
| 45 | TMT-B Time@Acute | 0.523 | 0.209 | 0.015 | -1.021 | 0.015 | 0.133 | -0.106 | | | |
| 46 | TMT-B Time@Asymptomatic | 0.229 | -0.835 | -0.250 | 0.100 | 0.433 | -0.025 | | | | |
| 47 | TMT-B Time@Baseline | 0.198 | -0.072 | -0.431 | 0.108 | 0.116 | -0.098 | | | | |
| 48 | Time Lost | 0.544 | -0.518 | -1.419 | 0.062 | | | | | | |

**Table B3**: Expanded scoring. This table shows the product between the trained weights $w_i$ and all the bins $\mathcal{B}^i$ for each variable $x_i$, which can be seen as a scorecard which associates a given occurrence (bin) with a score. The sum of all the occurrences $\sum_i w_i\mathcal{B}^i$ for a given athlete plus the bias term $w_0 = -1.093$ provide the log-odds ratio $\log \frac{\mathbb{P}(Y=1|\boldsymbol{x})}{\mathbb{P}(Y=0|\boldsymbol{x})}$.

| $i$ | Variable | $\exp(w_i\mathcal{B}_1^i)$ | $\exp(w_i\mathcal{B}_2^i)$ | $\exp(w_i\mathcal{B}_3^i)$ | $\exp(w_i\mathcal{B}_4^i)$ | $\exp(w_i\mathcal{B}_5^i)$ | $\exp(w_i\mathcal{B}_6^i)$ | $\exp(w_i\mathcal{B}_7^i)$ | $\exp(w_i\mathcal{B}_8^i)$ | $\exp(w_i\mathcal{B}_9^i)$ | $\exp(w_i\mathcal{B}_{10}^i)$ |
|---|---|---|---|---|---|---|---|---|---|---|---|
| 1 | BESS@Asymptomatic | 0.882 | 0.653 | 1.035 | 1.141 | 1.241 | 0.917 | | | | |
| 2 | BESS@Baseline | 2.066 | 0.790 | 0.731 | 0.385 | 1.398 | 2.230 | 0.561 | | | |
| 3 | BSI@Acute | 1.306 | 0.491 | 2.861 | 0.699 | | | | | | |
| 4 | CRT Difference Baseline Acute | 1.399 | 0.229 | 1.306 | 1.452 | 4.916 | 0.703 | | | | |
| 5 | CRT Difference Baseline RTA | 0.683 | 1.107 | 1.351 | 0.949 | | | | | | |
| 6 | CRT@Asymptomatic | 1.165 | 0.836 | 0.698 | 1.059 | 1.452 | 0.875 | | | | |
| 7 | HADS Depression | 1.875 | 1.007 | 0.939 | 0.841 | 0.356 | 0.790 | | | | |
| 8 | ImPACT Motor Speed Difference Baseline Asymptomatic | 1.326 | 1.100 | 0.984 | 0.617 | 1.423 | 0.938 | | | | |
| 9 | ImPACT Motor Speed@Baseline | 0.996 | 0.675 | 1.040 | 1.338 | 0.861 | | | | | |
| 10 | ImPACT Motor Speed@RTA | 0.600 | 0.404 | 0.726 | 1.014 | 1.299 | 1.264 | | | | |
| 11 | ImPACT Reaction Time Difference Baseline Asymptomatic | 1.009 | 0.919 | 0.686 | 1.009 | 1.114 | 1.219 | 1.736 | 0.949 | | |
| 12 | ImPACT Reaction Time Difference Baseline RTA | 1.365 | 0.619 | 0.821 | 0.961 | 1.079 | | | | | |
| 13 | ImPACT Reaction Time@Asymptomatic | 5.595 | 1.406 | 0.345 | 0.679 | 1.022 | 1.022 | | | | |
| 14 | ImPACT Reaction Time@RTA | 1.738 | 1.029 | 0.609 | 0.295 | 1.161 | 1.344 | | | | |
| 15 | ImPACT Verbal Memory Difference Baseline Acute | 0.808 | 1.018 | 1.096 | 1.831 | 1.198 | 1.034 | 0.881 | | | |
| 16 | ImPACT Verbal Memory Difference Baseline RTA | 0.271 | 1.213 | 2.186 | 0.967 | 0.683 | 1.105 | | | | |
| 17 | ImPACT Verbal Memory@RTA | 0.271 | 0.673 | 1.389 | 1.117 | 0.927 | 1.246 | | | | |
| 18 | ImPACT Visual Memory Difference Baseline Acute | 1.333 | 0.475 | 1.114 | 1.917 | 0.836 | | | | | |
| 19 | ImPACT Visual Memory@Acute | 1.011 | 0.665 | 1.056 | 1.184 | 1.194 | 0.833 | | | | |
| 20 | ImPACT Visual Memory@Baseline | 1.006 | 0.977 | 0.946 | 0.967 | 1.017 | 1.023 | 0.967 | | | |
| 21 | K-D Time Difference Baseline Acute | 1.182 | 0.946 | 1.035 | | | | | | | |
| 22 | K-D Time Difference Baseline Asymptomatic | 2.329 | 1.220 | 0.273 | 0.833 | 1.529 | 1.702 | 0.883 | | | |
| 23 | K-D Time Difference Baseline RTA | 1.615 | 0.415 | 0.975 | 0.917 | | | | | | |
| 24 | K-D Time@Acute | 0.643 | 1.217 | 2.902 | 0.456 | 0.828 | | | | | |
| 25 | K-D Time@Baseline | 0.825 | 1.192 | 3.542 | 0.323 | 0.795 | | | | | |
| 26 | K-D Time@RTA | 0.676 | 0.881 | 1.764 | 1.162 | 1.071 | 0.973 | 0.958 | | | |
| 27 | NPC@Acute | 1.128 | 0.399 | 1.936 | 0.905 | | | | | | |
| 28 | NPC@Baseline | 1.011 | 1.556 | 4.493 | 0.525 | 0.848 | | | | | |
| 29 | NPC@RTA | 0.987 | 1.185 | 0.987 | 0.981 | | | | | | |
| 30 | SAC@Asymptomatic | 3.044 | 1.333 | 1.084 | 0.774 | 0.951 | 0.866 | | | | |
| 31 | SCAT Severity Difference Baseline Asymptomatic | 2.859 | 1.401 | 1.248 | 0.553 | 0.600 | 1.075 | 0.816 | | | |
| 32 | SWLS | 0.321 | 1.052 | 1.064 | 6.621 | 1.192 | 1.045 | 0.623 | | | |
| 33 | Sport | 0.404 | 0.465 | 0.749 | 0.999 | 1.019 | 1.058 | 1.391 | 1.600 | 2.184 | 3.398 |
| 34 | TG DT Difference Baseline RTA | 0.656 | 1.854 | 0.729 | 1.011 | | | | | | |
| 35 | TG DT@Asymptomatic | 0.592 | 2.355 | 1.073 | 1.008 | | | | | | |
| 36 | TG DT@Baseline | 0.880 | 1.431 | 1.669 | 1.640 | 0.842 | 0.616 | 0.867 | | | |
| 37 | TG DT@RTA | 1.401 | 0.434 | 1.023 | 1.588 | 0.976 | | | | | |
| 38 | TG ST@Acute | 1.550 | 0.850 | 0.462 | 1.280 | 0.957 | | | | | |
| 39 | TG ST@Baseline | 1.457 | 0.989 | 0.412 | 2.879 | 0.724 | | | | | |
| 40 | TMT-A Time Difference Baseline Acute | 0.850 | 0.887 | 0.981 | 1.244 | 1.024 | | | | | |
| 41 | TMT-A Time Difference Baseline Asymptomatic | 2.361 | 0.628 | 0.220 | 1.023 | 1.218 | 0.973 | | | | |
| 42 | TMT-A Time@Acute | 1.641 | 0.280 | 0.350 | 1.031 | 1.729 | 3.148 | 0.810 | | | |
| 43 | TMT-A Time@Asymptomatic | 1.102 | 1.040 | 0.765 | 0.990 | | | | | | |
| 44 | TMT-A Time@Baseline | 2.108 | 1.367 | 1.213 | 0.421 | 0.982 | 1.210 | 0.856 | | | |
| 45 | TMT-B Time@Acute | 1.687 | 1.232 | 1.016 | 0.360 | 1.016 | 1.142 | 0.899 | | | |
| 46 | TMT-B Time@Asymptomatic | 1.258 | 0.434 | 0.779 | 1.106 | 1.542 | 0.976 | | | | |
| 47 | TMT-B Time@Baseline | 1.218 | 0.930 | 0.650 | 1.114 | 1.123 | 0.906 | | | | |
| 48 | Time Lost | 1.724 | 0.596 | 0.242 | 1.064 | | | | | | |

**Table B4**: Exponentiated expanded scoring. This table shows the exponentiation of the product between the trained weights $w_i$ and all the bins $\mathcal{B}^i$ for each variable $x_i$. We can interpret each value in this table as a factor that makes the prediction more or less likely to be a post-concussion MSK injury, assuming all the other variables involved in the computation are fixed.

| $i$ | Variable | $\lvert\mathcal{B}_1^i\rvert$ | $\lvert\mathcal{B}_2^i\rvert$ | $\lvert\mathcal{B}_3^i\rvert$ | $\lvert\mathcal{B}_4^i\rvert$ | $\lvert\mathcal{B}_5^i\rvert$ | $\lvert\mathcal{B}_6^i\rvert$ | $\lvert\mathcal{B}_7^i\rvert$ | $\lvert\mathcal{B}_8^i\rvert$ | $\lvert\mathcal{B}_9^i\rvert$ | $\lvert\mathcal{B}_{10}^i\rvert$ |
|---|---|---|---|---|---|---|---|---|---|---|---|
| 1 | BESS@Asymptomatic | 9 | 15 | 44 | 22 | 32 | 33 | | | | |
| 2 | BESS@Baseline | 8 | 14 | 9 | 19 | 60 | 21 | 24 | | | |
| 3 | BSI@Acute | 74 | 12 | 13 | 56 | | | | | | |
| 4 | CRT Difference Baseline Acute | 10 | 8 | 19 | 24 | 20 | 74 | | | | |
| 5 | CRT Difference Baseline RTA | 14 | 60 | 12 | 69 | | | | | | |
| 6 | CRT@Asymptomatic | 16 | 15 | 9 | 48 | 22 | 45 | | | | |
| 7 | HADS Depression | 42 | 37 | 10 | 23 | 10 | 33 | | | | |
| 8 | ImPACT Motor Speed Difference Baseline Asymptomatic | 8 | 51 | 18 | 11 | 9 | 58 | | | | |
| 9 | ImPACT Motor Speed@Baseline | 26 | 10 | 86 | 15 | 18 | | | | | |
| 10 | ImPACT Motor Speed@RTA | 10 | 8 | 27 | 16 | 25 | 69 | | | | |
| 11 | ImPACT Reaction Time Difference Baseline Asymptomatic | 8 | 23 | 8 | 16 | 19 | 15 | 8 | 58 | | |
| 12 | ImPACT Reaction Time Difference Baseline RTA | 9 | 9 | 8 | 52 | 77 | | | | | |
| 13 | ImPACT Reaction Time@Asymptomatic | 12 | 27 | 9 | 35 | 24 | 48 | | | | |
| 14 | ImPACT Reaction Time@RTA | 9 | 35 | 20 | 13 | 9 | 69 | | | | |
| 15 | ImPACT Verbal Memory Difference Baseline Acute | 13 | 19 | 22 | 11 | 19 | 14 | 57 | | | |
| 16 | ImPACT Verbal Memory Difference Baseline RTA | 8 | 10 | 8 | 43 | 9 | 77 | | | | |
| 17 | ImPACT Verbal Memory@RTA | 8 | 16 | 12 | 9 | 41 | 69 | | | | |
| 18 | ImPACT Visual Memory Difference Baseline Acute | 8 | 8 | 67 | 15 | 57 | | | | | |
| 19 | ImPACT Visual Memory@Acute | 8 | 10 | 17 | 57 | 17 | 46 | | | | |
| 20 | ImPACT Visual Memory@Baseline | 43 | 8 | 9 | 9 | 10 | 58 | 18 | | | |
| 21 | K-D Time Difference Baseline Acute | 8 | 75 | 72 | | | | | | | |
| 22 | K-D Time Difference Baseline Asymptomatic | 13 | 38 | 10 | 18 | 8 | 9 | 59 | | | |
| 23 | K-D Time Difference Baseline RTA | 34 | 8 | 46 | 67 | | | | | | |
| 24 | K-D Time@Acute | 10 | 65 | 15 | 8 | 57 | | | | | |
| 25 | K-D Time@Baseline | 11 | 85 | 13 | 14 | 32 | | | | | |
| 26 | K-D Time@RTA | 9 | 19 | 10 | 32 | 9 | 25 | 51 | | | |
| 27 | NPC@Acute | 47 | 10 | 21 | 77 | | | | | | |
| 28 | NPC@Baseline | 61 | 8 | 14 | 10 | 62 | | | | | |
| 29 | NPC@RTA | 33 | 13 | 33 | 76 | | | | | | |
| 30 | SAC@Asymptomatic | 11 | 18 | 20 | 29 | 45 | 32 | | | | |
| 31 | SCAT Severity Difference Baseline Asymptomatic | 8 | 18 | 45 | 12 | 9 | 14 | 49 | | | |
| 32 | SWLS | 10 | 30 | 11 | 16 | 21 | 38 | 29 | | | |
| 33 | Sport | 9 | 8 | 36 | 21 | 24 | 11 | 14 | 12 | 11 | 9 |
| 34 | TG DT Difference Baseline RTA | 10 | 14 | 11 | 120 | | | | | | |
| 35 | TG DT@Asymptomatic | 15 | 8 | 34 | 98 | | | | | | |
| 36 | TG DT@Baseline | 9 | 22 | 17 | 11 | 15 | 8 | 73 | | | |
| 37 | TG DT@RTA | 20 | 10 | 8 | 11 | 106 | | | | | |
| 38 | TG ST@Acute | 10 | 13 | 8 | 37 | 87 | | | | | |
| 39 | TG ST@Baseline | 58 | 26 | 10 | 9 | 52 | | | | | |
| 40 | TMT-A Time Difference Baseline Acute | 8 | 32 | 11 | 12 | 92 | | | | | |
| 41 | TMT-A Time Difference Baseline Asymptomatic | 20 | 10 | 8 | 16 | 21 | 80 | | | | |
| 42 | TMT-A Time@Acute | 17 | 8 | 12 | 8 | 21 | 20 | 69 | | | |
| 43 | TMT-A Time@Asymptomatic | 26 | 60 | 15 | 54 | | | | | | |
| 44 | TMT-A Time@Baseline | 8 | 8 | 27 | 13 | 18 | 37 | 44 | | | |
| 45 | TMT-B Time@Acute | 14 | 39 | 8 | 8 | 8 | 9 | 69 | | | |
| 46 | TMT-B Time@Asymptomatic | 39 | 12 | 15 | 23 | 12 | 54 | | | | |
| 47 | TMT-B Time@Baseline | 32 | 23 | 10 | 36 | 10 | 44 | | | | |
| 48 | Time Lost | 26 | 10 | 10 | 109 | | | | | | |

**Table B5**: Expanded counts. This table shows the number of athletes that are taken into account in each bin. The sum of each row amounts to the total number of athletes in the training set ($N = 155$).

### Appendix C Model Robustness

Assessing the robustness of predictive models is critical to ensure their reliability and applicability across diverse datasets and settings. This section describes the experiments we performed to evaluate the robustness of the predictive model generated by our methodology. That is, we execute a comprehensive resampling analysis with random stratified training/testing splits and evaluate the variability of model performance, feature selection, and feature importance.

#### C.1 Resampling Procedure and Model Training Scenarios

We generate 20 independent stratified resamples of the data set to ensure consistent outcome distributions between the training and testing splits, keeping the data proportions described in Section 2.4. This means that each training set contains 155 athletes and each test set comprises 39 athletes. During the resampling process, we discard splits where categorical variables introduce unseen categories in the test set, which only happens for rare cases on rare categories due to the 4 to 1 ratio of train to test.

#### C.2 Model Performance Analysis

To evaluate model performance, we analyze two scenarios for each data split: (a) executing the complete modeling pipeline, learning the WoE transformation, applying it to the training and testing sets, conducting feature selection, and training a logistic regression model with feature selection; and (b) executing the pipeline without feature selection, retaining the same 48 variables chosen by our proposed model. For both scenarios, performance metrics, including precision, recall, F1-score, AUROC, and average precision, are calculated for each trained model on its respective test set.

| **Approach** | **Metric** | **Mean** | **Std** | **Min** | **50%** | **Max** |
|---|---|---|---|---|---|---|
| Feature selection | $F_1^\star$ | 0.845 | 0.031 | 0.800 | 0.844 | 0.902 |
| | AP | 0.856 | 0.040 | 0.786 | 0.854 | 0.912 |
| | AUROC | 0.801 | 0.030 | 0.761 | 0.792 | 0.872 |
| | Precision@$F_1^\star$ | 0.786 | 0.061 | 0.667 | 0.797 | 0.900 |
| | Recall@$F_1^\star$ | 0.921 | 0.065 | 0.750 | 0.917 | 1.000 |
| Fixed variable | $F_1^\star$ | 0.841 | 0.031 | 0.792 | 0.841 | 0.898 |
| | AP | 0.848 | 0.042 | 0.765 | 0.841 | 0.930 |
| | AUROC | 0.792 | 0.046 | 0.686 | 0.796 | 0.892 |
| | Precision@$F_1^\star$ | 0.767 | 0.063 | 0.676 | 0.776 | 0.880 |
| | Recall@$F_1^\star$ | 0.938 | 0.055 | 0.792 | 0.958 | 1.000 |

**Table C6**: Performance metrics variablitity in key performance metrics (precision, recall, F1-score, ROC AUC, and average precision) across 20 resampled training/testing splits.

Table C6 highlights the variability in performance metrics across resamples. Models trained with a fixed set of variables exhibited slightly higher variability in performance compared to those utilizing feature selection, which demonstrated more consistent results. Generally, models with feature selection outperformed fixed-variable models, as the selection process identifies the most relevant predictors for each data split. However, the performance differences was relatively small, indicating that the 48 fixed variables are still effective predictors of post-concussion MSK injury risk. For models trained using feature selection, the average number of selected variables across all resamples was 47, ranging from a minimum of 41 to a maximum of 52, with a total of 112 distinct variables being selected throughout the process.

To complement the analysis of model performance, we compute performance curves (ROC and Precision-Recall) for predictions grouped by sports across resamples. For each trained model on a resampled training set, we collect the predictions made on the corresponding test set. Since the same athlete can appear in multiple test sets across the 20 splits, we average the probabilistic predictions for each athlete across all test set appearances. Once all model outputs are collected and the predictions for the same athletes are averaged, the data is segregated by the sport variable to generate the ROC and Precision-Recall curves shown in Figures C5 and C6.

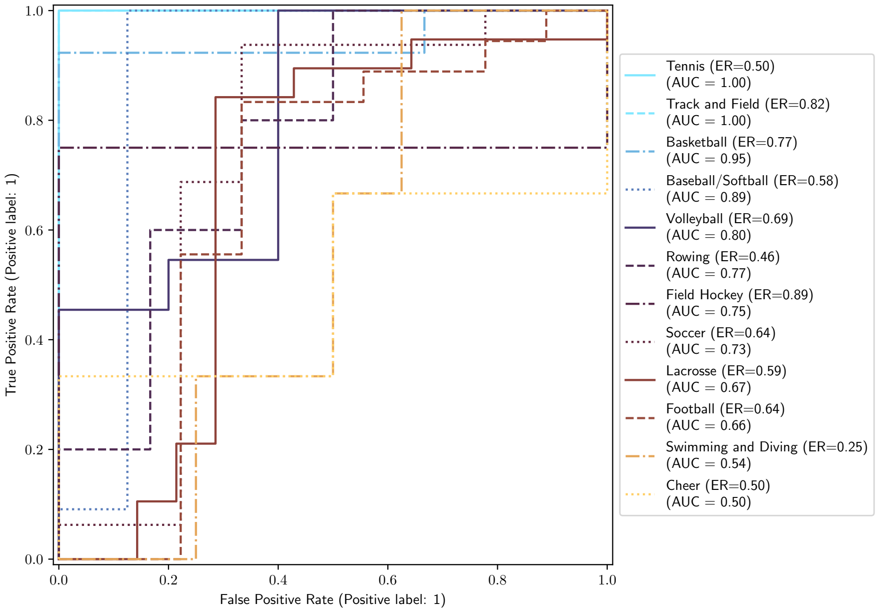


**Fig. C5**: ROC curve per sport. These curves illustrate the model's ability to differentiate between athletes who experienced a post-concussion musculoskeletal injury (MSK) and those who did not grouped by sports.

The performance for Tennis and Track and Field achieves near-perfect metrics, with both having an Area Under the Curve (AUC) and Average Precision (AP) values of 1.00. This indicates the model's exceptional ability to discriminate between high- and low-risk athletes in these sports. However, the small sample size for Tennis (only

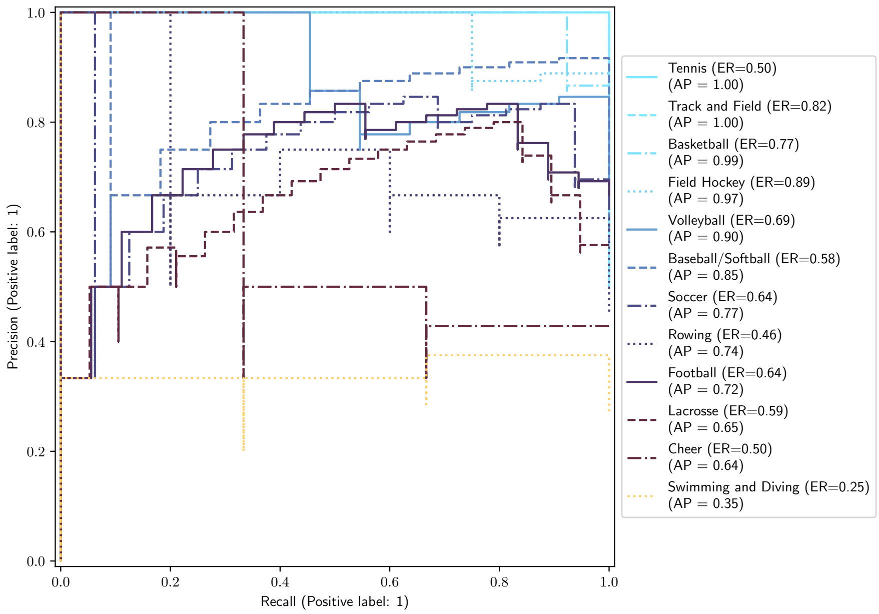


**Fig. C6**: AP curve per sport. These plots highlight the trade-off between precision (positive predictive value) and recall (sensitivity) in classifying athletes who are at risk of post-concussion MSK injuries grouped by sports.

four athletes) should be noted as a limitation, suggesting these results may be overly optimistic. Basketball, Volleyball, and Field Hockey also showed strong performance, with AP values ranging from 0.90 to 0.97 and AUCs from 0.80 to 0.95. These results demonstrate that the model captures the relevant risk patterns for these sports effectively. Sports like Baseball/Softball, Soccer, Rowing, and Football exhibited moderate predictive performance, with AP values between 0.72 and 0.85 and AUC values ranging from 0.66 to 0.89. Lacrosse had an AP value of 0.65 and an AUC of 0.67. These results indicate that, while the model can provide meaningful predictions, its discriminatory power is less robust in these sports compared to the previous ones. Cheer along with Swimming and Diving had the lowest predictive performance, with AP values ranging from 0.35 to 0.65 and AUCs between 0.50 and 0.54. This suggests that the model struggles to identify clear risk patterns for athletes in these sports, which involve unique aspects. Differences in performance between sports could be due to the varying relevance of predictive variables in predicting MSK injury in different athletic activities. After the WoE transformation, the risk is a generalized linear model, which cannot account for second-order interactions between sport and other variables, highlighting opportunities for model refinement.

#### C.3 Feature Selection Analysis

To assess feature selection consistency, we construct a variable selection incidence matrix (Figure C7) with counts of how many times each variable is included in a model. For longitudinal variables, we track their occurrences across all time points, ensuring each time point is counted only once, even if a variable appears in multiple forms (e.g., `TMT-A@Acute` and `TMT-A Difference Baseline Acute`). Additionally, we create a variable correlation matrix using the WoE-transformed variables from each training data set, and compute the average correlations to provide deeper insights into variable relationships.

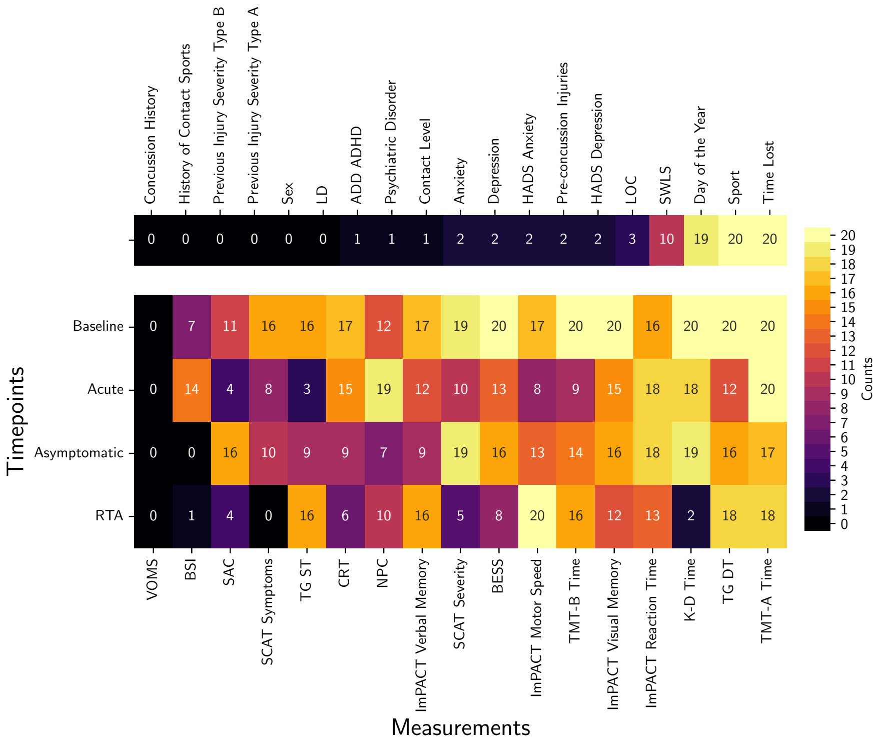


**Fig. C7**: Variable selection incidence. Heatmaps showing the number of times a variable was selected during 20 stratified resampling splits for static and longitudinal variables. The upper heatmap is for the 19 static variables measured at a single timepoint. The lower heatmap focuses on 17 longitudinal variables measured at four timepoints (Baseline, Acute, Asymptomatic, and RTA) for which a variable (a measurement at a time point) is considered selected if it is selected alone or as part of a difference between Baseline and one of the three other timepoints (excluding `VOMS` which was not involved in differences). Total number of variables is $135 = 19 + 17 \times 4 + 16 \times 3$.

The top heatmap in Figure C7 shows how frequently the variables that are measured only once, such as baseline demographics and clinical characteristics, are selected during the modeling process. Key variables like `Time Lost` and `Sport` are consistently selected in all 20 iterations, highlighting their strong predictive significance. In addition, the variable `Day of the Year` is frequently chosen, underscoring the importance of capturing seasonality-related patterns, as concussions towards the end of seasons may be associated with lower subsequent injury risk. `SWLS` is selected in half of the models. In contrast, variables such as `LOC` are selected less frequently.

The lower heatmap in Figure C7 illustrates the selection frequencies of longitudinal variables measured at four time points: Baseline, Acute, Asymptomatic, and Return-to-Activity (RTA). Baseline measurements generally appear highly relevant, as they are frequently selected, with the exception of `VOMS`. Variables such as `TMT-A Time` and `TG DT` are consistently chosen at all time points, emphasizing their critical importance in predicting the risk of musculoskeletal injury. Notably, certain variables show time-point-specific relevance. For example, `KD Test` is regularly chosen at Baseline, Acute, and Asymptomatic stages, but not at RTA. Similarly, `SCAT Symptoms` is often selected at Baseline and Asymptomatic stages, but is less prominent at the other timepoints. Another noteworthy observation is that `BSI` appears more relevant at the Acute time point compared to the Baseline, highlighting its temporal specificity in providing predictive information.

The average variable correlation matrix, shown in Figure C8, provides a more detailed view of the relationships between the variables in the dataset. The y-axis constitutes the top 50 variables derived from the multiple feature selection processes across resamples, while the x-axis represents the variables in our proposed model. The matrix is divided into four quadrants:

- **Upper Left Quadrant (symmetric):** Displays the correlations among the 31 variables that are part of both the top 50 and our proposed model. This quadrant highlights some of the correlation within the core predictive variables.
- **Lower Left Quadrant:** Shows the relationships between the remaining 19 top variables not included in our model and the other 31 variables in our model that are in the top 50. This area reveals high correlation among a few top variables in the model with those not selected.
- **Upper Right Quadrant:** Represents the relationships between the 31 top-selected variables and the 17 variables in our model that are not part of the top 50.
- **Lower Right Quadrant:** Illustrates the correlations between the 17 model-specific variables that are not part of the top 50 and the 19 remaining top variables outside our model.

The strong positive correlation between `K-D Time@Baseline` and `K-D Time Difference Baseline RTA` implies that the risk associated to athletes' performance on the King-Devick test at baseline is correlated to the risk associated to the change observed between baseline and RTA. Also, a noticeable positive correlation exists between `ImPACT Motor Speed@Asymptomatic` and `ImPACT Motor Speed@RTA`, indicating correlated risks associated to the motor speed measurements at the asymptomatic and return-to-activity time periods.

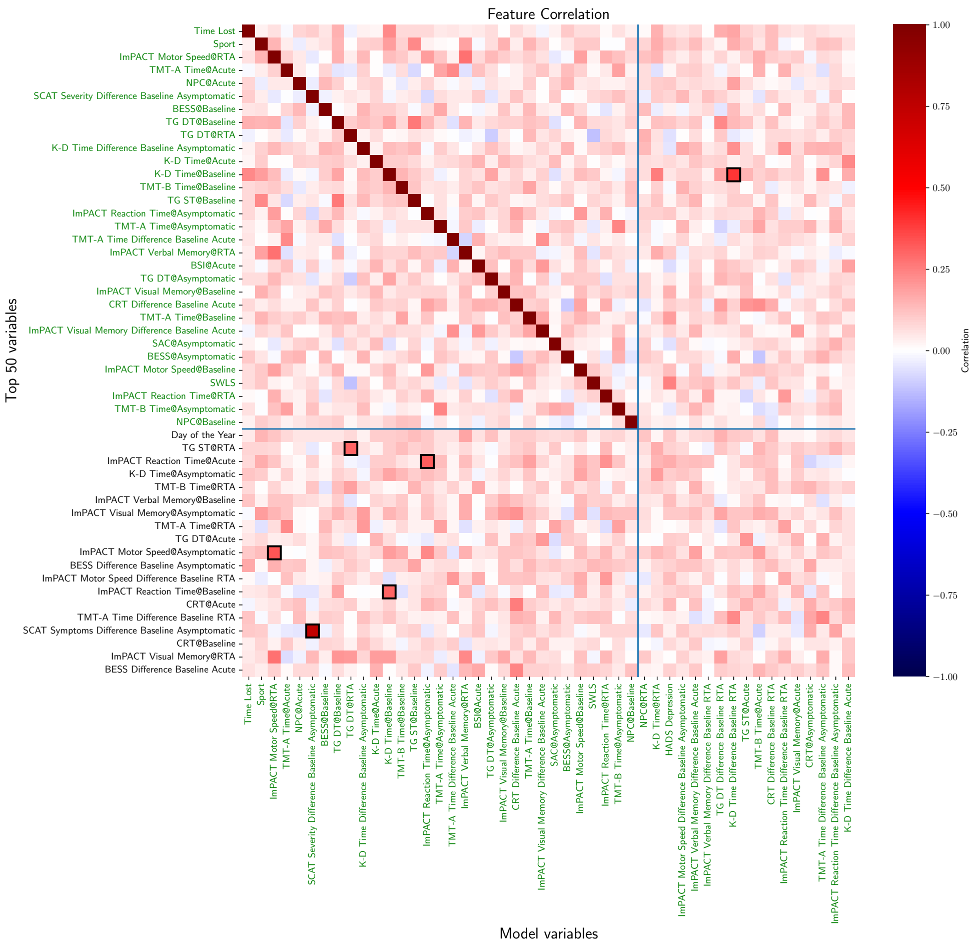

**Fig. C8**: Average transformed variable correlation. This matrix depicts the correlation between WoE-transformed variables across 20 resampled models. The names of variables in green correspond to the variables that belong to our model.

The average correlation matrix helps explain why the total number of non-repeated variables selected across all models (112) is significantly higher than the average number of selected variables (47), since with high correlation, certain pairs of variables are interchangeable. For example, `TG ST@RTA` and `TG DT@RTA`, and `SCAT Symptoms Difference Baseline Asymptomatic` and `SCAT Severity Difference Baseline Asymptomatic` are strongly correlated and can provide similar information. This explains why feature selection algorithms may alternate between these variables across resampling iterations, reinforcing the robustness of our approach despite variations in selected features. The presence of correlated variables also explains why only 31 of the 48 selected in our proposed model are present among the most selected variables.

#### C.4 Feature Importance Analysis

To analyze feature importance, we compute SHAP values for each model and compute the variability of SHAP values across splits. Analyzing the SHAP value box plots provides a comprehensive understanding of feature importance. `Sport` consistently emerges as a highly influential predictor, highlighting the significance of considering the specific sport in assessing the risk of post-concussion MSK injury due to the inherent differences in injury risk between sports. Variables related to the Trail Making Test, such as `TMT-A Time@Acute` and `TMT-A Time Difference Baseline Acute`, consistently demonstrate high importance. This underlines the value of including cognitive performance measures in risk prediction models. Variables with high mean SHAP values, such as `K-D Time Difference Baseline Asymptomatic` and `K-D Time@Acute` also show notable contributions to the predictions, emphasizing the information this test provide at different time points.

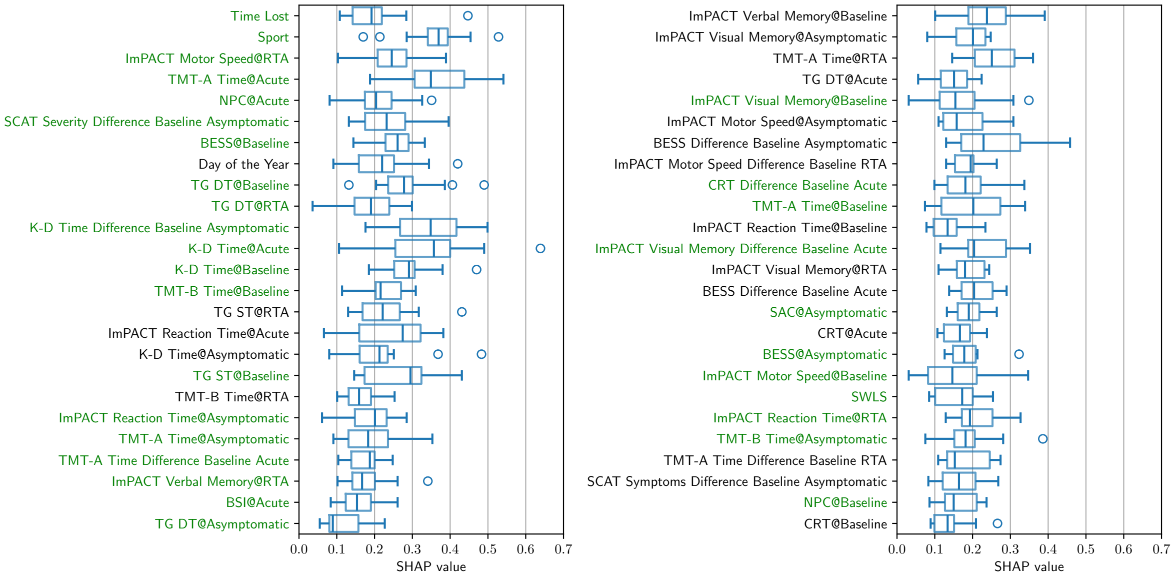


**Fig. C9**: SHAP values variability. Box plots of SHAP values for the top 50 most frequently selected variables across models trained on 20 resampled training sets.

Several consistencies emerge when comparing the resampled data analysis with the findings of our proposed approach. The importance of `Sport` and clinical assessment variables like the `TMT-A Time` and `ImPACT Motor Speed` remains consistent in both analyses. This reinforces the robustness of these predictors in identifying at-risk athletes. The frequent co-selection of variables measuring the same concept at different time points, such as `TMT-A Time@Acute` and `TMT-A Time Difference Baseline Acute`, suggests that considering both the absolute values and the changes from baseline is crucial for accurate risk assessment.

However, some anomalies also merit attention. Although the original study found `Time Lost` to be a significant predictor, and the variable is selected across all splits, in terms of its mean SHAP value across resampled splits appears to be typically less important, with the SHAP value of the original split being an outlier. This discrepancy could be attributed to variations in variable selection due to complementary variables making it less important.

#### C.5 Evaluation of the Impact of Sports-related variables

Distributional shifts, such as variations in athlete demographics or institutional practices, can occur when replicating the study in different settings. It is challenging to anticipate potential shifts using data from a single institution; nonetheless, we consider forming a model when the critical sport-specific predictors are absent, which would enable the model to be applied to athletes from sports not in the current dataset. Low variability in performance metrics across resamples would suggest that the modeling approach is robust and likely to perform consistently across institutions with different athlete populations.

We exclude the variables `Sport`, `Contact Level`, and `Day of the Year` since the latter two both carry some information associated to `Sport`. Among these, `Sport` and `Day of the Year` are highly relevant in the original feature selection process and are identified as highly predictive based on its average SHAP value and high selection frequency across resamples.

We consider two scenarios for this evaluation for each resample: (1) Sport-related variables are removed entirely from the dataset prior to executing the modeling pipeline; and (2) the feature selection results of scenario (b) in Section C.2 are retained, but the coefficients of sport-related variables were forced to zero during logistic regression model fitting. The results of these two scenarios are presented in Table C7, capturing the model's performance across 20 resamples. We observe that the exclusion of sports-related generates a reduced performance, considering our proposed model's metrics. However, the stable performance across resamples suggest that the remaining variables in the dataset provide sufficient predictive power to compensate for their absence. These findings emphasize the adaptability of our approach and its potential for replication in different institutional contexts, even when key sport-specific predictors are unavailable or vary significantly across datasets.

| Approach | Metric | Mean | Std | Min | 50% | Max |
|---|---|---|---|---|---|---|
| Sport-related variables removed from feature selection process | $F_1^\star$ | 0.832 | 0.025 | 0.787 | 0.832 | 0.885 |
| | AP | 0.842 | 0.041 | 0.776 | 0.834 | 0.911 |
| | AUROC | 0.777 | 0.046 | 0.681 | 0.768 | 0.861 |
| | Precision@$F_1^\star$ | 0.763 | 0.058 | 0.649 | 0.768 | 0.870 |
| | Recall@$F_1^\star$ | 0.923 | 0.068 | 0.833 | 0.938 | 1.000 |
| Sport-related variables removed from feature-selection-based models (scenario (b), Section C.2) | $F_1^\star$ | 0.832 | 0.027 | 0.774 | 0.838 | 0.880 |
| | AP | 0.845 | 0.042 | 0.773 | 0.839 | 0.912 |
| | AUROC | 0.786 | 0.033 | 0.719 | 0.785 | 0.850 |
| | Precision@$F_1^\star$ | 0.780 | 0.055 | 0.632 | 0.786 | 0.870 |
| | Recall@$F_1^\star$ | 0.900 | 0.064 | 0.750 | 0.917 | 1.000 |
| All variables with feature selection | $F_1^\star$ | 0.845 | 0.031 | 0.800 | 0.844 | 0.902 |
| | AP | 0.856 | 0.040 | 0.786 | 0.854 | 0.912 |
| | AUROC | 0.801 | 0.030 | 0.761 | 0.792 | 0.872 |
| | Precision@$F_1^\star$ | 0.786 | 0.061 | 0.667 | 0.797 | 0.900 |
| | Recall@$F_1^\star$ | 0.921 | 0.065 | 0.750 | 0.917 | 1.000 |
| Fixed variables from original model | $F_1^\star$ | 0.841 | 0.031 | 0.792 | 0.841 | 0.898 |
| | AP | 0.848 | 0.042 | 0.765 | 0.841 | 0.930 |
| | AUROC | 0.792 | 0.046 | 0.686 | 0.796 | 0.892 |
| | Precision@$F_1^\star$ | 0.767 | 0.063 | 0.676 | 0.776 | 0.880 |
| | Recall@$F_1^\star$ | 0.938 | 0.055 | 0.792 | 0.958 | 1.000 |

**Table C7**: Performance metrics for two scenarios where sport-related variables were excluded, along with results from Table C6. Metrics include $F_1^\star$, average precision (AP), AUROC, precision, and recall, with results averaged over 20 resamples. Both scenarios demonstrate robust model performance, with minor trade-offs in precision and recall.
